# Genetic liability for cardiovascular disease, physical activity, and mortality – findings from The Finnish Twin Cohort

**DOI:** 10.1101/2023.04.28.23289250

**Authors:** Laura Joensuu, Katja Waller, Anna Kankaanpää, Teemu Palviainen, Jaakko Kaprio, Elina Sillanpää

## Abstract

**Background:** An overall healthy lifestyle may mitigate the risk of all-cause and cardiovascular disease (CVD) mortality despite the inherited risk for CVD. We assessed if similar phenomenon is observed independently with physical activity (PA).

**Methods:** In a prospective older Finnish Twin Cohort, genetic liability for coronary heart disease, systolic and diastolic blood pressure was estimated with polygenic risk scores (PRSs) derived from the Pan UK Biobank (N≈400,000) and >1,000,000 genetic variants, and leisure-time PA longitudinally with validated and structured questionnaires three times during 1975–1990 among 4,897 participants aged 33–60-years (54.3% women). Interactions of PRSs and PA with mortality and the independent main effects of different PA metrics with mortality were evaluated with Cox proportional hazards models. A co-twin control design with 180 monozygotic twin pairs discordant for physical activity was used for causal inference.

**Results:** During the 17.4-y (mean) follow-up (85,136 person-years), 1,195 participants died with 389 CVD deaths. Although the favorable associations of an overall healthy lifestyle with mortality risk reduction replicated in the cohort, no interactions or significant independent main effects were observed with any of the assessed PA metrics. Adherence to World Health Organizations PA guidelines or vigorous PA during the 15-year observational period did not decrease mortality in more active twins compared to their less active identical twin brother or sister. The findings did not vary with genetic CVD risk.

**Conclusions:** No evidence was found that PA mitigates inherited CVD risk or is causally associated with mortality challenging some of our current understanding.

## Introduction

Cardiovascular diseases (CVD) remain persistently as leading causes of premature death worldwide, posing a considerable burden to both public and personal health.^1^ While modifiable lifestyle factors contribute to the onset of the disease,^2,3^ the role of genetic factors is not negligible.^4^ Family history has long been established as an independent risk factor for common CVDs,^5^ with heritability estimates of 40–60% based on twin studies.^6^

Previous landmark studies have promisingly shown that by adhering to an overall healthy lifestyle (regular physical activity, no obesity, no smoking, and healthy diet) the risk of CVD incidence could be mitigated despite possessing a risky genotype.^2,3^ Similar findings have emerged with CVD mortality.^7^ In this study, we evaluated the independent role of physical activity (PA) in risk mitigation while genetic liability for CVD is assessed directly from the genome. PA is commonly promoted by public health authorities to increase life expectancy,^8^ with consistent supporting epidemiological evidence among different demographics and with different PA metrics.^9–14^ We investigated if PA during adult years mitigates the risk of mortality despite the genetic liability for CVD, and evaluated evidence for causality of this association.

## Methods

### Study oversight

The older Finnish Twin Cohort (FTC) study is a prospective cohort launched in 1975 and described in detail elsewhere.^15^ In brief, all same-sex twins born before 1958 and living in Finland were invited to the study with a 84.4% response rate. The study provides a unique 45-year follow-up resource for studying the role of lifestyle factors in health. Despite the twin structure, mortality in the FTC does not differ from the Finnish population.^16^ The study was funded with grants from Academy of Finland, and several foundations (Juho Vainio foundation, Päivikki and Sakari Sohlberg foundation, and Sigrid Juselius foundation), and the Wellcome Trust Sanger Institute, the Broad Institute, ENGAGE – European Network for Genetic and Genomic Epidemiology); the funders did not influence or involved in any phases of the study. The FTC study was evaluated by ethics committees of the University of Helsinki (113/E3/01 and 346/E0/05), Helsinki University Central Hospital (136/E3/01, 01/2011, 270/ 13/03/01/2008, and 154/13/03/00/2011). The authors assume responsibility for the accuracy and completeness of the protocol, data, and analyses, fidelity for their reporting, and complying with the Declaration of Helsinki and guidelines of the Finnish Advisory Board on Research Integrity.

### Study design

In this study, we utilised a genotyped sub-sample of the FTC (See Figures S2 and S3 for flow charts). The participants answered to structured and validated questionnaires in 1975, 1981 and 1990 (see Figure S4 for study design). Later, participants were invited to DNA-sampling, taking place at individual time points between 1993 and 2017 (Figure S5). Their vital status was assessed in 2020. The follow-up began from DNA-sampling and ended at death, migration, or end of the follow-up on December 31, 2020.

### Study procedures

In the first phase, we aimed to replicate the previous studies and assessed if an overall healthy lifestyle mitigates the risk of mortality despite genetic liability for CVD in the FTC cohort. In the second phase, we assessed the interactions between PA and genetic liability for CVDs, and independent main effects of PA with mortality utilising different PA metrics. In the third phase, we utilised a co-twin control design with monozygotic (MZ) twin pairs for causal inference. In exploratory analyses we assessed if the genetic predisposition to CVD associates with PA levels.

### Outcomes

All-cause mortality and CVD mortality were assessed. Dates and causes of death by December 31^st^ 2020 were retrieved from the Population Register Centre of Finland, and Statistics Finland. All-cause mortality included all events leading to death. CVD mortality was based on Statistics Finland national time series classification (54 categories). Categories 27–30 represent CVD mortality, including ischemic heart diseases (corresponding ICD-10 codes (I20–I25), other forms of heart diseases except rheumatic and alcoholism-based (I30– I425, I427–I52), cerebrovascular diseases (I60–I69), hypertensive, pulmonary heart disease, disease of pulmonary circulation and disease of arteries, arterioles and capillaries, veins, lymphatic vessels, lymph nodes, and other unspecified disorders of the circulatory system (I100–I15, I26–I28, I70–I99).^17,18^

### Genetic liability for cardiovascular diseases, and physical activity

#### Polygenic risk scores

Polygenic risk scores (PRSs) summarise the genome-wide information between single-nucleotide polymorphisms (SNPs) and a certain phenotype into a single variable that quantifies person’s genetic liability to a disease or a trait.^19^ PRSs were calculated for three CVDs; coronary heart disease (CHD), systolic blood pressure (SBP) and diastolic blood pressure (DBP). After evaluating a few candidates for PRSs (Tables S2-S4), PRSs based on genome-wide association data from Pan-UK Biobank (restricted to participants with European ancestry) and the SBayesR method were selected. Pan-UK Biobank data included 396,663–419,724 participants (Table S1). We used SBayesR pipeline to construct the PRSs based on 1,005,933–1,006,472 genetic variants. More information related to construction of the PRSs is provided in the Supplemental Appendix, Chapter 1.1.

#### Physical activity

PA during leisure-time was assessed with structured and validated questionnaires in 1975, 1981 and 1990.^15,20,21^ For utilized questions, see Supplemental Appendix Chapter 2.3. Several PA metrics were formed. A metabolic equivalent of task (MET) in hours per week (h · week^-1^) was calculated as a product of reported intensity, duration and frequency of activity.^20^ Meeting the World Health Organisations PA guidelines for aerobic activity was defined as a minimum of 7.5 MET h · week^-1^,^11^ and calculated based on the mean MET h · week^-1^ in 1975–1990. Additional metrics were the continuous mean MET h · week^-1^ in 1975–1990, meeting the PA guidelines in all three measurement points (1975, 1981 and 1990), and adhering to vigorous PA in all three measurement points (1975, 1981 and 1990). Vigorous physical activity was considered as performing at least intermittent jogging.

### Covariates

Age, sex, educational attainment, body mass index, smoking, fruit and vegetable consumption, and alcohol consumption were assessed by questionnaires. The last known and/or most comprehensive data was used (1981 or 1990). Additionally, health status, i.e., self-reported symptoms, physician-diagnosed diseases and utilized medications were assessed by questionnaires and from national health register data between 1971 and 1983. Ten principal components of ancestry were used to adjust for any genetic stratification which may occur in the Finnish population. For further details related to covariates, see Supplementary Appendix, Chapter 2.3.

### Statistical analysis

The description and rationale of the statistical procedures, along with utilized codes, are provided in the Statistical Analysis Plan (SAP), which was written after accessing the data and prior to final analyses. A convenience sample was used, with all assumed and eligible twins in Finland being contacted to participate in 1975. In post hoc analysis, the main effects analyses indicate sufficient 80% power to detect the expected 29% decrease in mortality, while the power in the co-twin control design may be limited (SAP Chapter 3.2). A complete case approach was selected despite potential bias to firmly retain the twin structure in the data. Decision was supported by analyses where the complete case sample did not differ from the full sample by PA at baseline (Table S5).

After examining proportional hazard assumptions with Shoenfeld and Martingale residuals (Supplemental Appendix, Chapter 2.2), outcomes were analysed with Cox Proportional Hazards Model. In the first phase, the main effects of different combinations of different lifestyles (described in detail in Table S7) and genetic disease risk categories (sample-specific low (<33.3%), intermediate (33.3–66.6%), or high genetic risk for CVD (> 66.6%)) were compared against mortality, with participants with high genetic risk and unhealthy lifestyles utilized as reference category. In the second phase, the interactions between PRSs and PA were examined (PRS × PA) to evaluate the potential moderator effect. Additionally, the independent main effects of PA and PRSs with mortality were assessed. Models were adjusted for covariates.

In the third phase, the within-pair differences in mortality were assessed among MZ twin pairs discordant for their PA levels. In a co-twin control design the other twin is exposed to an environmental factor (e.g., meeting selected PA criteria) while the other twin is not. This difference in their exposure is evaluated against difference in their survival. The co-twin control design in MZ twins is considered to be an elegant study design for causal inference in epidemiological studies, as many potential confounding factors are shared among identical twins (genotype and childhood environments, with many characteristics in adulthood).^22,23^

Models are by design controlled for genetic factors and early life environmental factors. Models with highest number of twin pairs were additionally adjusted for main lifestyle risk factors; BMI, and smoking status.

In exploratory analyses, the association between genetic liability for CVD and different lifestyle factors were evaluated with linear and multinomial logistic regression. Throughout the study, all analyses were conducted separately for the three different genetic liabilities, with results related to genetic liability for CHD presented in the main text. As diseases are a potential source for reverse causality, sensitivity analyses were conducted with only apparently healthy participants. Across all analyses the twin structure of the data was acknowledged. Data was assumed to be missing at random. Statistical significance was set at P < 0.01 after Bonferroni correction for multiple testing (for three different CVDs) and 99% Confidence Interval reported. Analyses were performed with Stata/IC 16.0.

## Results

### Characteristics of the study sample

The descriptives of the eligible 4,897 participants are presented in Table 1 by their survival status at the end of follow-up. During the 17.4-y (mean) follow-up (85,136 person-years) 1,195 participants died with 389 CV deaths.

**Table 1.** Subject characteristics according to the latest known information by survival status at the end of follow-up.

|  | Alive |  | Deceased |  |
| --- | --- | --- | --- | --- |
|  | Mean (SD) |  | Mean (SD) |  |
|  | N | or % | N | or % |
| **N=4,897** |  |  |  |  |
| **Characteristic** |  |  |  |  |
| Age (years) | 3,702 | 44.8 (7.2) | 1,195 | 51.3 (6.9) |
| Male (%) | 1,540 | 41.6 | 699 | 58.5 |
| Female (%) | 2,162 | 58.4 | 496 | 41.5 |
| Height (cm) | 3,702 | 168.3 (8.8) | 1,195 | 169.3 (8.8) |
| Weight (kg) | 3,702 | 70.1 (13.1) | 1,195 | 74.4 (13.4) |
| BMI (kg · m<sup>-2</sup>) | 3,702 | 24.6 (3.7) | 1,195 | 25.9 (3.9) |
| **Educational level<sup>a</sup>** |  |  |  |  |
| Low (%) | 1,223 | 33.0 | 577 | 48.3 |
| Middle (%) | 1,827 | 49.4 | 502 | 42.0 |
| High (%) | 652 | 17.6 | 116 | 9.7 |
| **Health status<sup>a</sup>** |  |  |  |  |
| Not healthy (%) | 484 | 13.1 | 268 | 22.4 |
| Healthy (%) | 3,218 | 86.9 | 927 | 77.6 |
| **Smoking** |  |  |  |  |
| Never (%) | 1,830 | 49.4 | 475 | 39.8 |
| Occasional (%) | 114 | 3.1 | 35 | 2.9 |
| Former (%) | 925 | 25.0 | 316 | 26.4 |
| Light (%) | 212 | 5.7 | 54 | 4.5 |
| Medium (%) | 362 | 9.8 | 135 | 11.3 |
| Heavy (%) | 259 | 7.0 | 180 | 15.1 |
| **Fruit and vegetable consumption per day<sup>a</sup>** |  |  |  |  |
| Not once (%) | 237 | 6.4 | 120 | 10.0 |
| Once or twice (%) | 3,071 | 83.0 | 993 | 83.1 |
| Three times or more (%) | 394 | 10.6 | 82 | 6.9 |
| **Alcohol consumption** |  |  |  |  |
| Lifetime abstainer (%) | 275 | 7.4 | 106 | 8.9 |
| Former (%) | 42 | 1.1 | 14 | 1.2 |
| Occasional (%) | 224 | 6.1 | 74 | 6.2 |
| Low (%) | 2,878 | 77.7 | 814 | 68.1 |
| Medium (%) | 199 | 5.4 | 115 | 9.6 |
| High (%) | 62 | 1.7 | 46 | 3.9 |
| Very high (%) | 22 | 0.6 | 26 | 2.2 |
| **Physical activity** |  |  |  |  |
| Mean PA 1975–1990 (MET h · week<sup>-1</sup>) | 3,702 | 19.1 (15.9) | 1,195 | 17.0 (14.1) |
| Meeting PA guidelines (avg MET h · week<sup>-1</sup> 1975–1990) | 2,983 | 80.6 | 893 | 74.7 |

Values are means and standard deviations for continues variables and proportions of all participants for others. Data from 1990 if not else specified; a,Data from 1981; BMI, Body mass index; PA, Physical activity; MET, Metabolic equivalents; Meeting PA guidelines, average of MET h · week<sup>-1</sup> during 1975–1990 ≥ 7.5.

### Validation and replication

PRSs indicating genetic liability to CVDs indicated valid mortality risk in the FTC. In crude models, one standard deviation increment in PRSs was associated with 6% to 7% and 13% to 17% increased risk in all-cause and CV mortality, respectively (Table S2). These associations persisted after adjustments in all-cause mortality for PRS for DPB, and in CVD mortality for all PRSs (Tables 2 and 3 for CHD, Tables S9–S12 for others).

**Table 2.**
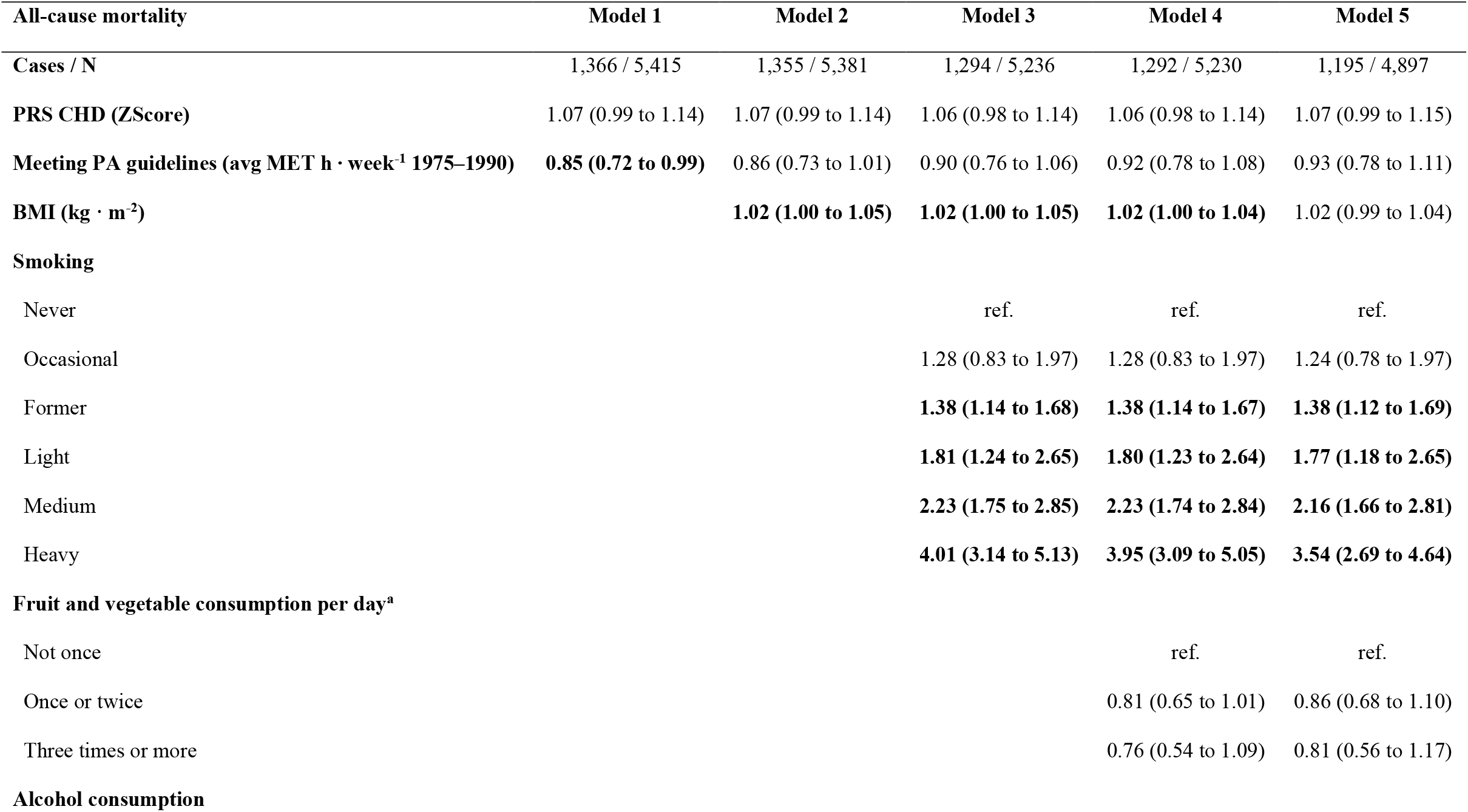
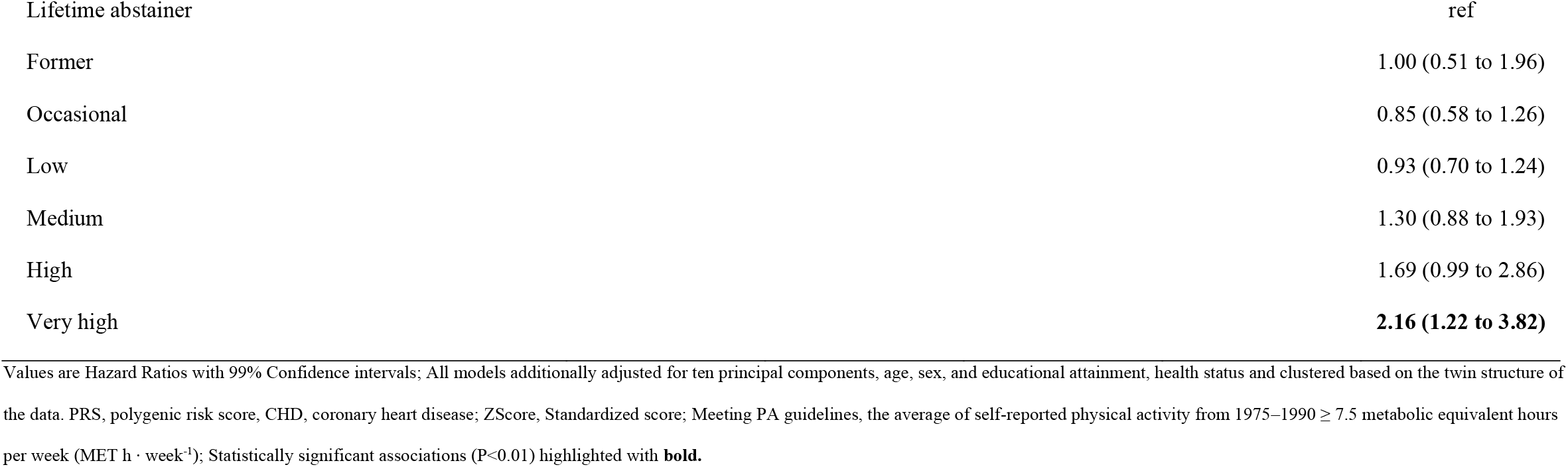
Associations of polygenic risk scores for coronary heart disease and physical activity with all-cause mortality after step-by-step adjustments for other healthy lifestyle factors.

**Table 3.**
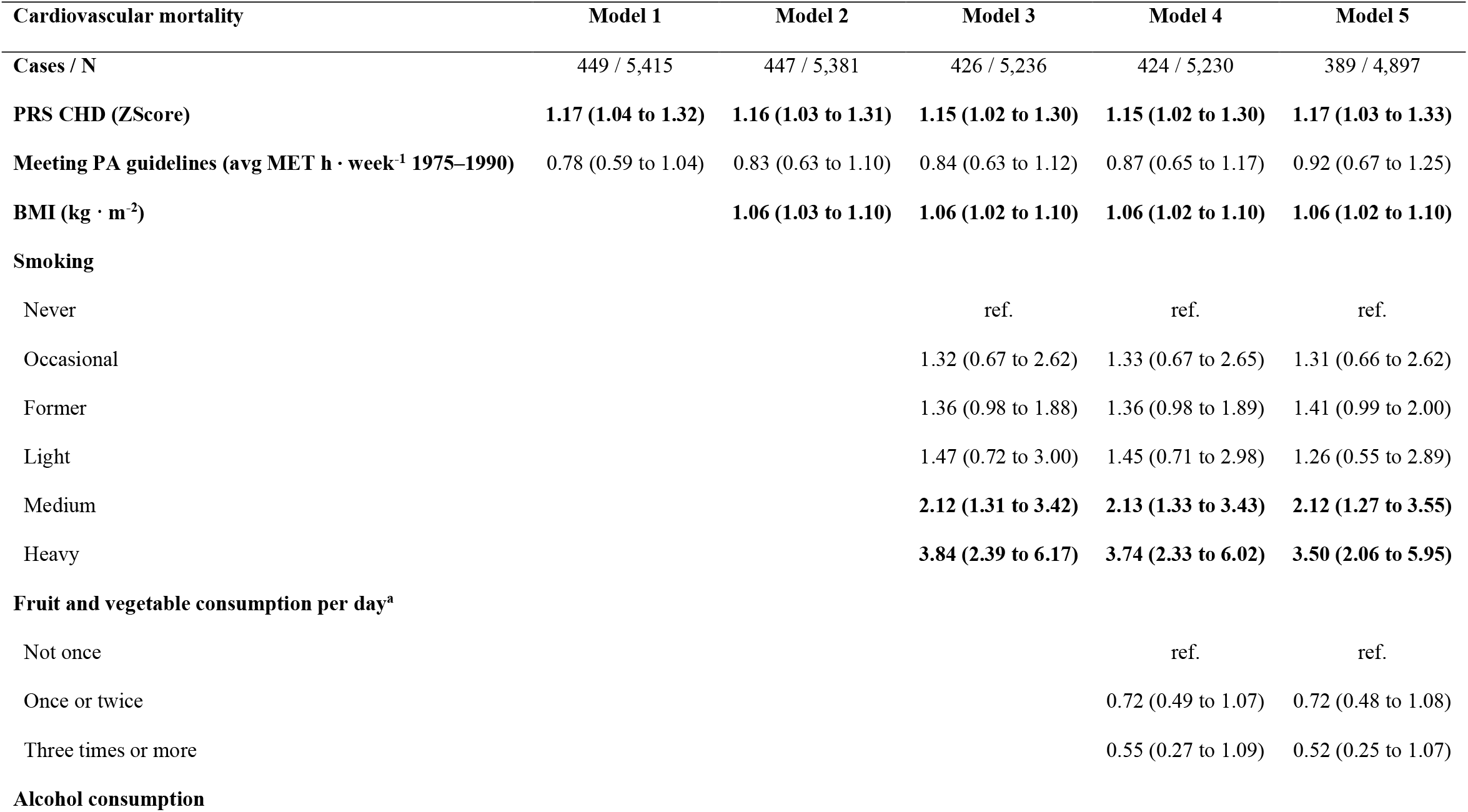
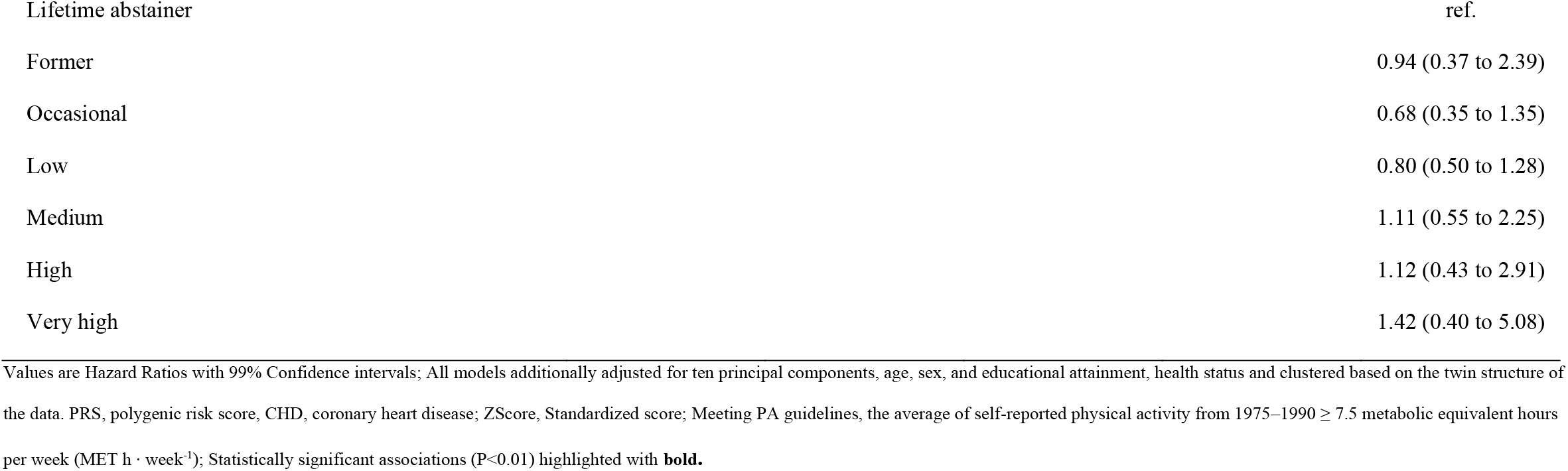
Associations of polygenic risk scores for coronary heart disease and physical activity with cardiovascular mortality after step-by-step adjustments for other healthy lifestyle factors.

The favourable associations of an overall healthy lifestyle replicated in the FTC and was associated with a 43% to 61% lower risk of all-cause and 57% to 76% CVD mortality, except in participants with high genetic liability for CHD (Figures S6 and S7).

### Interactions and independent main effects of PA with mortality

No interactions between different PA metrics and PRSs were observed with all-cause or CVD mortality (Table S8). The independent favorable associations of meeting the PA guidelines with mortality gradually attenuated when accounting for other healthy lifestyles from Hazard Ratio 0.85 (99% Confidence interval 0.72 to 0.99) to 0.93 (0.78 to 1.11) in all-cause mortality and from 0.78 (0.59 to 1.04) to 0.92 (0.67 to 1.25) in CVD mortality (Tables 2 and 3 for CHD, and S9–S12 for others). Of covariates, smoking was consistently associated with higher risk of mortality, BMI only with CVD mortality and high alcohol consumption only with all-cause mortality. No statistically significant associations were observed with any of the additionally evaluated PA metrics (Tables S13–S18), or in sensitivity analyses with only healthy participants (Tables S19–S21).

### Causal relationship between PA and mortality

The twin pairs in co-twin control design did not differ from one another by other characteristics than their PA (Figure 1). Meeting the PA guidelines did not statistically significantly decrease the risk of all-cause 0.74 (0.37 to 1.48), or CVD mortality 0.39 (0.10 to 1.46) in within-pair comparison between more and the less active twins (Table 5). Findings were similar across genetic liabilities for different CVDs and genetic disease risk categories (Table 5, S22 and S23), in healthy twin pairs (Tables S24 and S25) and among the three evaluated PA discordances (Tables S26–S29).

**Figure 1.**
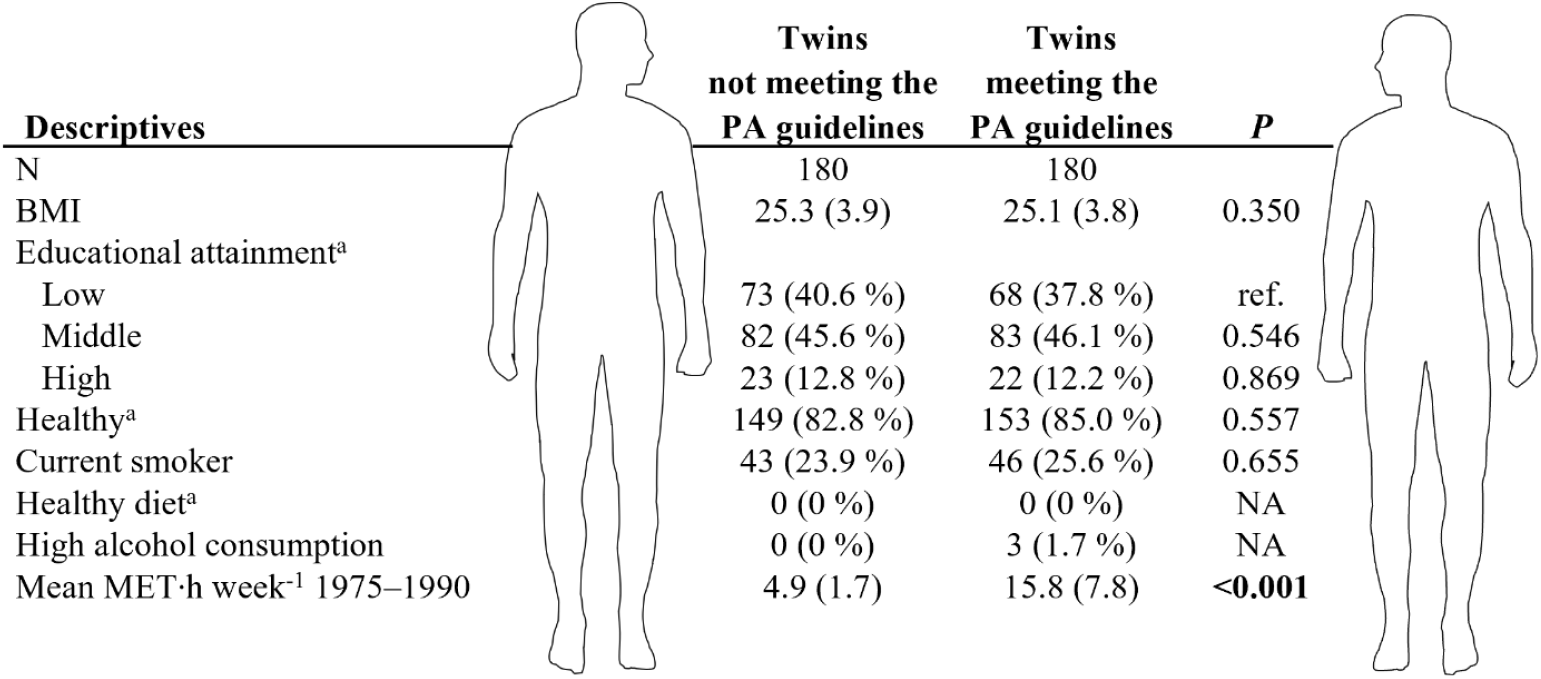
Descriptives of the monozygotic twin pairs, in which one twin meets the PA guidelines and the other twin does not. Values are means and standard deviations or proportions (%). Meeting PA guidelines, the average of self-reported physical activity from 1975–1990 ≥ 7.5 metabolic equivalent hours per week (MET h · week^-1^); Data from 1990 if not else specified; a, Data from 1981; BMI, Body mass index; *P*, statistical difference between the twins who meet the PA guidelines or not; NA, Not applicable due low number of twins. Statistically significant associations (P<0.01) highlighted with **bold**.

**Table 5.**
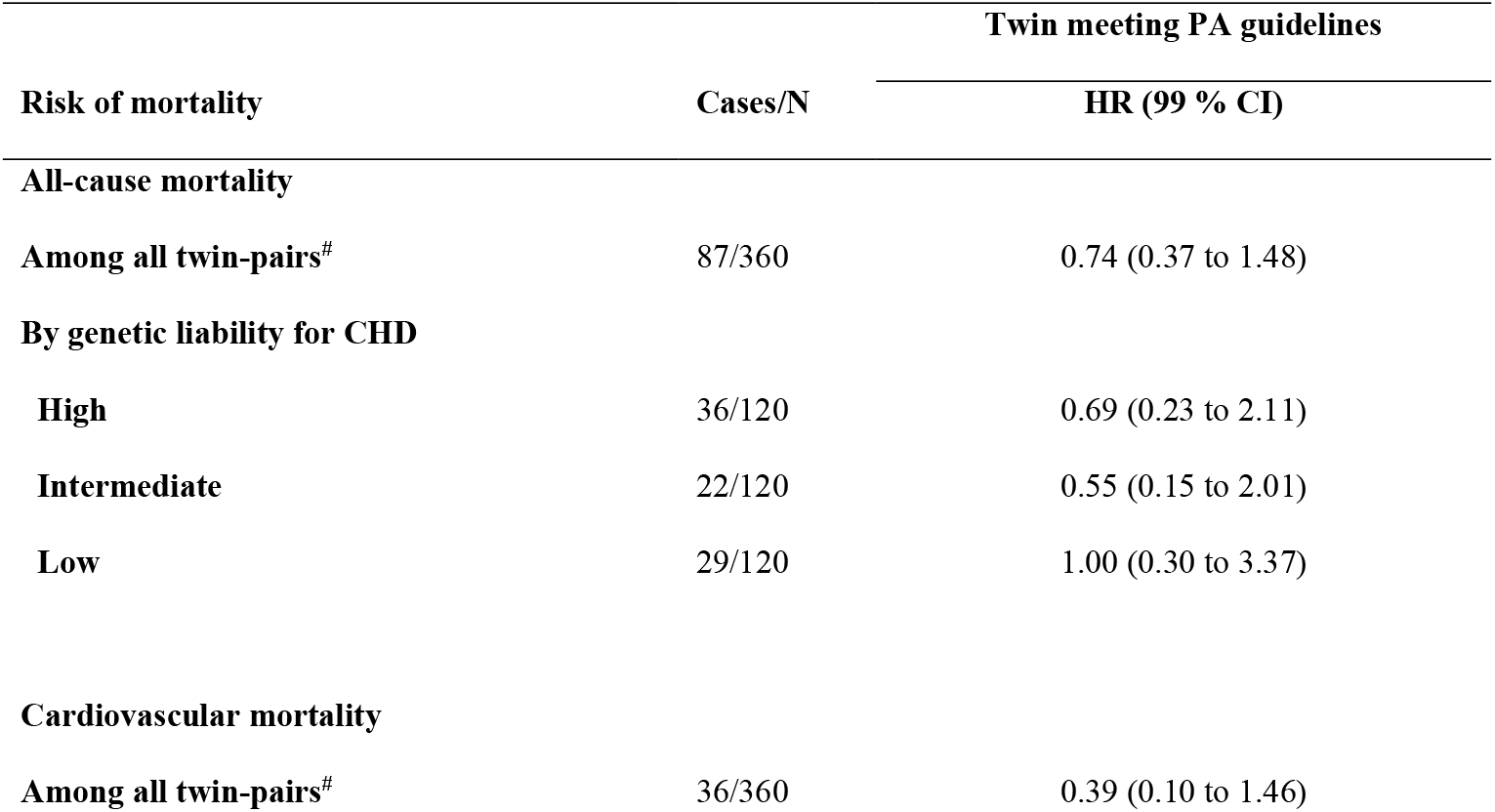
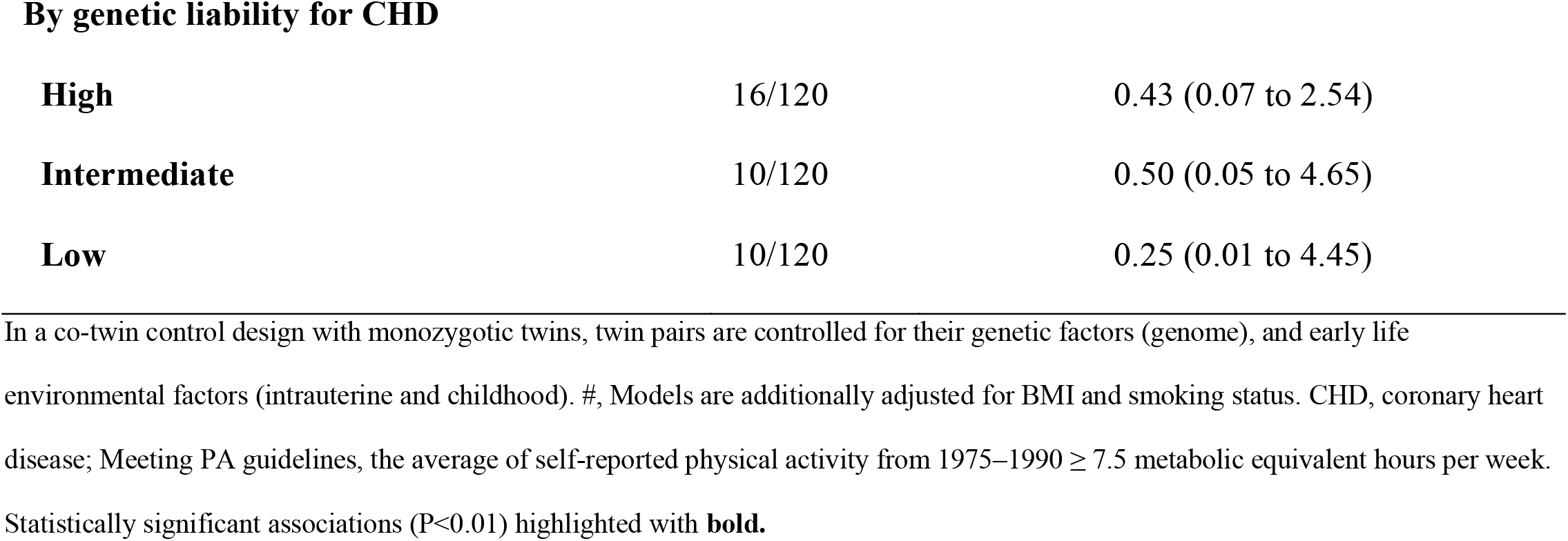
Within-pair risk of mortality in more active twins compared to their less active twin for 360 twins from 180 discordant pairs

| Risk of mortality | Cases/N | Twin meeting PA guidelines |
| --- | --- | --- |
|  |  | HR (99 % CI) |
| All-cause mortality |  |  |
| Among all twin-pairs<sup>#</sup> | 87/360 | 0.74 (0.37 to 1.48) |
| By genetic liability for CHD |  |  |
| High | 36/120 | 0.69 (0.23 to 2.11) |
| Intermediate | 22/120 | 0.55 (0.15 to 2.01) |
| Low | 29/120 | 1.00 (0.30 to 3.37) |
| Cardiovascular mortality |  |  |
| Among all twin-pairs<sup>#</sup> | 36/360 | 0.39 (0.10 to 1.46) |
| **By genetic liability for CHD** |  |  |
| **High** | 16/120 | 0.43 (0.07 to 2.54) |
| **Intermediate** | 10/120 | 0.50 (0.05 to 4.65) |
| **Low** | 10/120 | 0.25 (0.01 to 4.45) |

In a co-twin control design with monozygotic twins, twin pairs are controlled for their genetic factors (genome), and early life environmental factors (intrauterine and childhood). #, Models are additionally adjusted for BMI and smoking status. CHD, coronary heart disease; Meeting PA guidelines, the average of self-reported physical activity from 1975–1990 $\geq 7.5$ metabolic equivalent hours per week. Statistically significant associations ( $P < 0.01$ ) highlighted with **bold**.

### Exploratory analyses

The exploratory analyses did not suggest that higher genetic liability for CVDs predisposes to lower PA levels. Interestingly, higher genetic liability for SBP and DBP, but not for CHD, was associated with higher BMI: regression coefficient 0.24 (99% Confidence Interval 0.09 to 0.38), 0.27 (0.12 to 0.41) and 0.12 (−0.02 to 0.26), respectively (Table S30).

## Discussion

In this study, we evaluated the role of PA in reducing mortality risk of genetic liability for CVD. Our study supported the previous findings that adherence to an overall healthy lifestyle attenuates the risk of all-cause and CVD mortality despite the inherited risk for CVD.^7^ However, after extensive evaluations, we did not find any independent favorable associations between PA and mortality. These findings question the potential to mitigate the adverse outcomes of inherited risk of CVD with PA.

The main observations are highlighted and critically discussed. Firstly, the phenomenon of positive correlations between an overall healthy lifestyle, favorable health outcomes, and reduced mortality despite genetic liability to CVD is reliably replicated. However, our findings underline that not all lifestyle factors contribute equally, and the role of PA is limited. Interestingly, this interpretation is supported by the seminal studies but overshadowed by emphasis on the overall healthy lifestyle concept.^2,3^ We recommend to carefully consider if overall lifestyle scores provide information that sufficiently supports the development of health promotion strategies.

Secondly, our study found no evidence of interactions between PA and genetic risk in relation to mortality. Although the existing literature is limited, our novel findings suggest that physical activity may not moderate the association between inherited CVD risk and mortality.

Thirdly, in our sample, the independent main effects of PA were attenuated in a step-wise manner when adjusting for other healthy lifestyles. Physically active individuals often possess also other healthy lifestyles,^24^ and this clustering might cause unobserved confounding if not appropriately considered. It is also important to acknowledge that although the main risk factors (smoking, BMI) may be most relevant, the associations may further attenuate with additional adjustments.

Finally, the twins who were more active during the extensive 15-year observational period did not live longer compared to their less active cotwin in identical twin pairs. Findings were similar with different PA metrics (meeting PA guidelines or not, adhering to vigorous PA or not), and with different genetic disease risk levels for different CVDs. Assessing causality between PA and mortality has been shown to be methodologically challenging,^25^ with our study being no exception. However, our study is in line with previous findings from randomized controlled trials (RCTs), animal studies, and Mendelian randomization studies where no convincing evidence for causal relationship between PA and mortality has been observed.^26–28^

We acknowledge that self-reported metrics are prone to biases, and data was collected during an era with potentially limited generalizability to modern lifestyles. Data lost during the long observational period reduced the number of subjects in complete cases analysis, potentially introducing bias. Larger sample sizes or PA discordances might have strengthened some of the observed associations.

### “Physical activity might not bring more years to life, but more life to years”

The above mentioned quote from professor Urho Kujala summarizes the public health message of this study. This study does not refute well-established findings from RCTs and the biological rationale that support the role of exercise and PA in primary and secondary CVD risk modification.^29,30^ Notably, previous findings from the FTC show that more active MZ twins manifest more favorable tissue and metabolic health alongside with better physical fitness compared to their less active co-twins, supporting the findings from RCTs.^22^ However, the discrepancy between the systematic observational findings and persistent inability to confirm causality between PA and mortality requires further thought. Plausible explanations are multifactorial; potential bias in observational studies may be induced by healthy exerciser bias, unobserved and residual confounding, reverse causation, and genetic predisposition for both lesser physical activity and increased disease risk.^26,31–33^ Hence, physical activity may predominantly reflect one’s health and fitness status by conveying the underlying biological milieu rather than causally reduce mortality. The limited independent role of PA is evident in identical twin pairs, where higher engagement in PA does not associate with longer life expectancy.

In conclusion, no evidence was found that PA mitigates the inherited CVD disease risk or is causally associated with mortality. The benefits of an overall lifestyle were explained mainly by lesser smoking and healthy BMI. These critical findings provide novel information for future health policies.

## Supporting information

Study Protocol and Statistical Analysis Plan

## Data Availability

All data produced in the present study are available upon reasonable request to the authors.

