## Supplementary Appendix for "Genetic liability for cardiovascular disease, physical activity, and mortality – findings from The Finnish Twin Cohort"

### 1. Data preprocessing

#### 1.1 Polygenic risk scores

##### 1.1.1 Base data

The Pan-UK biobank dataset was used as the base data. The Pan-UK Biobank is an open access database with hundreds of thousands of individuals' genetic data paired to electronic health records and survey measures.[1] We selected cardiovascular disease (CVD) data related to coronary heart disease (CHD), systolic blood pressure (SBP) and diastolic blood pressure (DBP) for further analyses (Table S1).

Table S1. Descriptives of the base data for polygenic risk scores.

| Source (Phenocode) | N (Cases/Controls)[a] | wget | SNP heritability $H^2$ (SE)[a] | intercept |
|---|---|---|---|---|
| **Ischemic Heart Diseases** | | | | |
| Pan-UK Biobank (411) | 419,724 (37,672/38,2052) | wget https://pan-ukb-us-east-1.s3.amazonaws.com/sumstats_flat_files/phecode-411-both_sexes.tsv.bgz | 0.0329 (0.0024) | 1.0585 |
| **Systolic Blood Pressure (SBP)** | | | | |
| Pan-UK Biobank (4080) | 396,663 | wget https://pan-ukb-us-east-1.s3.amazonaws.com/sumstats_flat_files/continuous-4080-both_sexes-irnt.tsv.bgz | 0.1296 (0.0054) | 1.2736 |
| **Diastolic Blood Pressure (DBP)** | | | | |
| Pan-UK Biobank (4079) | 396,667 | wget https://pan-ukb-us-east-1.s3.amazonaws.com/sumstats_flat_files/continuous-4079-both_sexes-irnt.tsv.bgz | 0.1226 (0.0056) | 1.2606 |

[a], Participants with European ancestry

1.1.2 SBayesR pipeline

A Bayesian multiple regression approach based on genome-wide base data was used to calculate the individual genetic disease risk.[2] Re-weighted summary statistics restricted to HapMap3 variants were formed based on the summary statistics of the Pan-UK Biobank. The constructed summary statistics included data from 1,005,933–1,006,472 genetic variants and were used to calculate the individual polygenic risk scores for the Finnish Twin Cohort participants. The detailed pipeline is described in detail elsewhere.[3]

1.1.3 PRS validation

Polygenic risk scores (PRSs) calculated by the SBayesR pipeline fitted well and predicted higher risk of mortality in the Finnish Twin Cohort (FTC) target data. All PRSs followed a normal distribution (Figure S1) and were statistically significantly associated with higher risk of all-cause (Hazard Ratio (HR) from 1.06 to 1.07 and CVD mortality (HR: 1.13 to 1.17) (Table S2).

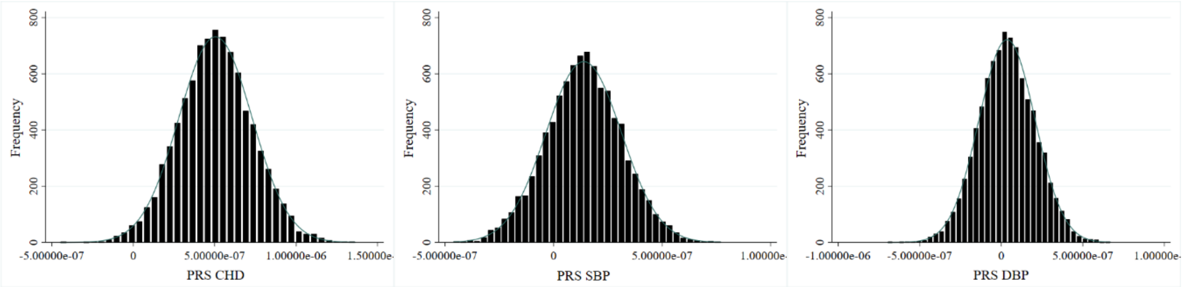

Figure S1. Distribution of the polygenic risk scores (PRS) for coronary heart disease (CHD), systolic blood pressure (SBP), and diastolic blood pressure (DBP) in the Finnish Twin Cohort target data set.

Table S2. Associations between polygenic risk scores and mortality in crude models.

| Variable | Hazard Ratio (95% Confidence Interval) | |
|---|---|---|
| | All-cause mortality | CVD mortality |
| PRS CHD SBayesR | **1.06 (1.02 to 1.10)** | **1.13 (1.06 to 1.20)** |
| PRS SBP SBayesR | **1.07 (1.03 to 1.11)** | **1.17 (1.10 to 1.25)** |
| PRS DBP SBayesR | **1.07 (1.03 to 1.11)** | **1.15 (1.08 to 1.22)** |

PRS, standardized polygenic risk scores; CHD, Coronary Heart Disease; SBP, Systolic Blood Pressure; DBP, Diastolic Blood Pressure; SBayesR, Bayesian multiple regression approach utilizing summary statistics from genome-wide association studies. Results indicate increase in mortality risk by one standard deviation increase in PRS. Models adjusted for ten principal components and clustered based on family number. Statistically significant associations highlighted with **bold**.

*Alternative PRSs*

As alternatives to the SBayesR method and the Pan-UK Biobank base data, other summary statistics and computational methods were considered. PRSs for SBP and DBP were formed from the Pan UK Biobank base data with alternative Bayesian approach taking account the linkage disequilibrium between each variant.[4,5] In addition, PRS for CHD was constructed by based on 49,310 single nucleotide polymorphism (SNPs) from CARDIoGRAMplusC4D Consortium meta-analysis and thinning the SNPs by linkage disequilibrium (LD) thresholds.[5]

Alternative PRSs performed also well and were statistically significantly associated with higher risk of all-cause (5.7–9.3%) and CVD mortality (11.8–18.0%) (Table S3). However, the SBayesR method and Pan-UK Biobank base data were selected for coherent approach. All evaluated PRSs correlated significantly with each other (Table S4), with correlations of 0.76 for systolic and diastolic blood pressure PRS alternatives with SBayesR-based PRS, but only 0.30 for the CHD PRSs potentially due differences in the base data and methods.

Table S3. Associations between alternative polygenic risk scores and mortality in crude models.

| Variable | Hazard Ratio (95% Confidence Interval) | |
|---|---|---|
| | All-cause mortality | CVD mortality |
| PRS CHD alternative | **1.06 (1.02–1.10)** | **1.12 (1.05–1.19)** |
| PRS SBP alternative | **1.07 (1.03–1.12)** | **1.16 (1.09–1.24)** |
| PRS DBP alternative | **1.09 (1.05–1.14)** | **1.18 (1.11–1.24)** |

CHD, Coronary Heart Disease; SBP, Systolic Blood Pressure; DBP, Diastolic Blood Pressure. Models adjusted for ten principal components and clustered based on family number. Statistically significant associations (P<0.05) highlighted with **bold**.

Table S4. Pearson correlations between different polygenic risk scores.

| N=8,838–8,856 | PRS CHD alternative | PRS SBP alternative | PRS DBP alternative | PRS CHD SBayesR | PRS SBP SBayesR | PRS DBP SBayesR |
|---|---|---|---|---|---|---|
| PRS CHD alternative | -- | | | | | |
| PRS SBP alternative | **0.132** | -- | | | | |
| PRS DBP alternative | **0.116** | **0.699** | -- | | | |
| PRS CHD SBayesR | **0.296** | **0.139** | **0.093** | -- | | |
| PRS SBP SBayesR | **0.146** | **0.760** | **0.509** | **0.142** | -- | |
| PRS DBP SBayesR | **0.118** | **0.514** | **0.762** | **0.082** | **0.653** | -- |

CHD, Coronary Heart Disease; SBP, Systolic Blood Pressure; DBP, Diastolic Blood Pressure; alternative, alternative methods were formed with an inhouse script (CHD) and a Bayesian approach taking account the linkage disequilibrium between each variant (SBP and DBP); SBayesR, Bayesian multiple regression approach utilizing summary statistics from genome-wide association studies. Models adjusted for ten principal components and clustered based on family number. Statistically significant (P<0.05) associations highlighted with **bold**.

##### 1.2 Genotyping, quality control and imputation

Genotyping in the UK-Biobank was performed with customized array and imputed to Haplotype Reference Consortium panel, yielding ~7 million SNPs (imputation quality score (INFO) ≥ 0.1 and MAF ≥ 1%).[6] Genotyping in the FTC was performed with Illumina Human610-Quad v1.0 B, Human670-QuadCustom v1.0 A, Illumina HumanCoreExome (12 v1.0 A, 12 v1.1 A, 24 v1.0 A, 24 v1.1 A, 24 v1.2 A), and Affymetrix FinnGen Axiom arrays and imputed to Haplotype Reference Consortium release 1.1 reference panel.[7]

### 2. Supplementary methods

The data was inspected for distribution and potential outliers. Potential moderator effect of sex in the association between physical activity and mortality was tested, but the interaction term was found non-significant (p=0.990). Therefore, the analyses were conducted for both sexes combined. In this document, individual analyses indicate to analyses conducted for the whole cohort, while pairwise analyses indicate causality assessment with the co-twin control design.

#### 2.1 Missing data

Participant flow charts are illustrated in Figures S2 and S3 with information of missing data.

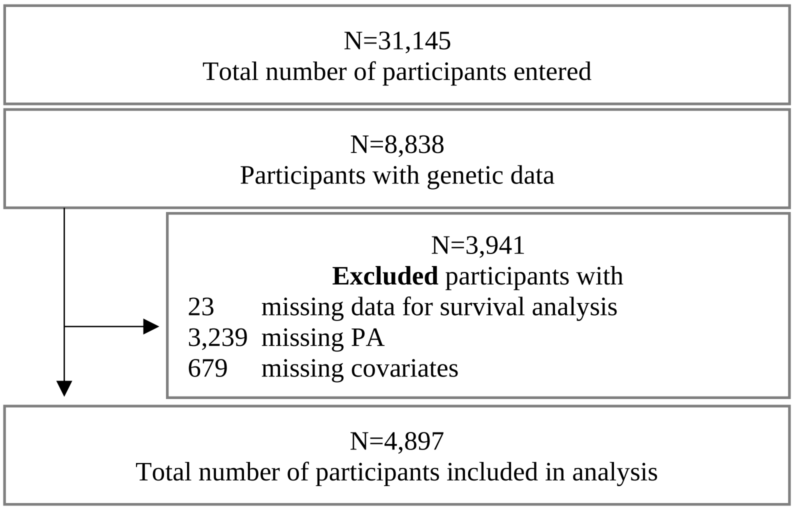

Figure S2. Participant flow chart of individual analyses

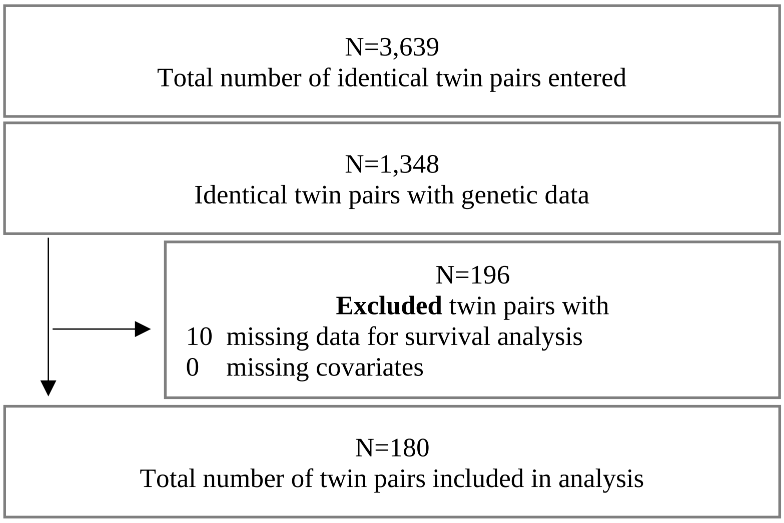

Figure S3. Participant flow chart of pairwise analyses

Subject characteristics at baseline for full FTC sample and the sample included in the final analysis (complete cases) are presented in Table S5. The differences between the samples were analysed with linear regression for continuous variables, logistic regression for binary, and multinominal regression for categorical variables and clustered based on the twin structure of the data. Participants of complete case analyses had more favorable lifestyles and better health compared to the full sample at baseline. The sample was younger, with lower BMI, and more likely healthy. Participants also had higher educational attainment and consumed alcohol more likely low levels, and less likely occasionally, compared to being lifetime abstainer. However, no differences were observed in physical activity, smoking or diet between full sample and complete case analysis sample at baseline.

Table S5. Subject characteristics at baseline in participants with FTC full sample and in the complete case sample.

| | **FTC full sample** | | **Final analysis sample (Complete cases)** | | |
|---|---|---|---|---|---|
| | **N** | **Mean (SD) or %** | **N** | **Mean (SD) or %** | ***P*** |
| **Characteristic** | | | | | |
| Age (years) | 8,838 | 34.0 (9.7) | 4,897 | 31.4 (7.7) | **<0.001** |
| Male | 4,159 | 47.1 | 2,239 | 45.7 | 0.013 |
| Height (cm) | 8,458 | 168.3 (8.8) | 4,884 | 168.6 (8.7) | **0.001** |
| Weight (kg) | 8,443 | 66.4 (12.0) | 4,876 | 65.6 (12.0) | **<0.001** |
| BMI (kg · $m^{-2}$) | 8,417 | 23.4 (3.3) | 4,865 | 23.0 (3.1) | **<0.001** |
| **Educational attainment**[a] | 8,046 | | 4,897 | | |
| Low (%) | 3,366 | 41.8 | 1,800 | 36.8 | ref. |
| Middle (%) | 3,573 | 44.4 | 2,329 | 47.6 | **<0.001** |
| High (%) | 1,107 | 13.8 | 768 | 15.7 | **<0.001** |
| **Health status**[a] | 8,838 | | 4,897 | | |
| Healthy | 6,372 | 72.1 | 4,145 | 84.6 | **<0.001** |

| | | | | | |
|---|---|---|---|---|---|
| **Smoking** | 8,467 | | 4,895 | | |
| Never (%) | 4,039 | 47.7 | 2,323 | 47.5 | ref. |
| Occasional (%) | 255 | 3.0 | 158 | 3.2 | 0.167 |
| Former (%) | 1,521 | 18.0 | 859 | 17.5 | 0.511 |
| Light (%) | 709 | 8.4 | 442 | 9.0 | 0.020 |
| Medium (%) | 1,225 | 14.5 | 715 | 14.6 | 0.613 |
| Heavy (%) | 718 | 8.5 | 398 | 8.1 | 0.320 |
| **Fruit and vegetable consumption per day**[a] | 8,255 | | 4,897 | | |
| Not once (%) | 651 | 7.9 | 357 | 7.3 | ref. |
| Once or twice (%) | 6,821 | 82.6 | 4,064 | 83.0 | 0.020 |
| Three times or more (%) | 783 | 9.5 | 476 | 9.7 | 0.026 |
| **Alcohol consumption** | 8,481 | | 4,897 | | |
| Lifetime abstainer (%) | 1,145 | 13.5 | 568 | 11.6 | ref. |
| Former (%) | 70 | 0.8 | 31 | 0.6 | 0.396 |
| Occasional (%) | 550 | 6.5 | 234 | 4.8 | **0.008** |
| Low (%) | 6,192 | 73.0 | 3,772 | 77.0 | **<0.001** |
| Medium (%) | 333 | 3.9 | 188 | 3.8 | 0.031 |
| High (%) | 137 | 1.6 | 78 | 1.6 | 0.116 |
| Very high (%) | 54 | 0.6 | 26 | 0.5 | 0.835 |
| **Physical activity** | | | | | |
| PA 1975 (MET h · week$^{-1}$) | 8,453 | 15.6 (19.0) | 4,897 | 15.6 (18.8) | 0.925 |
| Meeting PA guidelines 1975 (%) | 5,120 | 60.6 | 3,000 | 61.3 | 0.136 |

Differences between participants with missing data and complete cases were calculated with linear, logistic, or multinominal regression and clustered based on the twin structure of the data. Values are means and standard deviations for continues variables and cases with proportions for others. Data from 1975 if not else specified. a,Data from 1981; BMI, Body mass index; PA, Physical activity; MET, Metabolic equivalents. Statistically significant associations (P<0.01) highlighted with **bold.**

#### 2.2 Cox proportional hazards model

Time to an event, i.e., survival analysis, and the effect of covariates on the survival time are common interests in health sciences. Cox proportional hazard model is a method by which the effect of multiple variables to the survival time can be evaluated. Cox proportional hazard model does not require data to be normally distributed and can utilize both continuous and binary variables. Cox proportional hazards model rely on two main assumptions: 1) The hazards are proportional over time, and 2) The relationship between log hazards and covariates are linear. We tested these assumptions by Schoenfeld and Martingale residuals. The polygenic risk scores and main physical activity variable did not violate the proportional hazards (Table S6). In visual inspection the Locally Weighted Scatterplot Smoothing (lowess smooth) was linear and continuous covariates age and BMI did not violate the assumption.

Table S6. Results of proportional-hazards assumption test.

| Variable | Global test probability | |
|---|---|---|
| | All-cause mortality | CVD mortality |
| PRS CHD SBayesR | 0.639 | 0.383 |
| PRS SBP SBayesR | 0.428 | 0.408 |
| PRS DBP SBayesR | 0.419 | 0.332 |
| Meeting PA guidelines avg MET (h · week$^{-1}$) 1975–1990 | 0.858 | 0.733 |

CHD, Coronary Heart Disease; SBP, Systolic Blood Pressure; DBP, Diastolic Blood Pressure; SBayesR, Bayesian multiple regression approach utilizing summary statistics from genome-wide association studies.

#### 2.3 Supplementary methods

*Study design*

Study design is illustrated in Figure S4. PA questionnaires were filled 1975, 1981 and 1990. Follow-up started with DNA-sampling and ended December 31st, 2020.

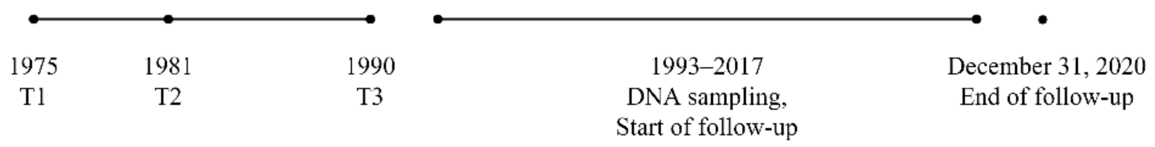

Figure S4. Study design

*Entry date*

In the Cox proportional hazards model, entry date i.e., the start of the follow-up was set to time of DNA-sampling to avoid immortal bias. DNA-sampling times are illustrated in Figure S5.

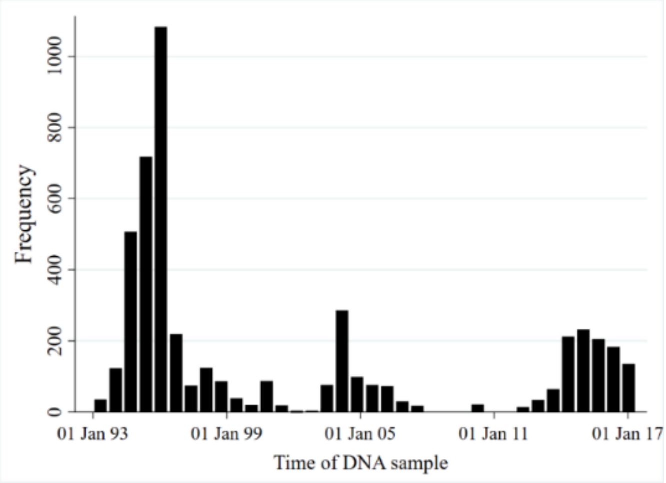

Figure S5. Dates of DNA sampling in the study sample.

*Physical activity questionnaire*

Original questionnaires are available in https://wiki.helsinki.fi/display/twineng/Older+cohort. Translations and specifications have been provided by Ph.D Katja Waller.[8] The 1975 and 1981 questionnaires were identical and shown below. The 1990 questionnaire differed slightly and included commuting to the MET score.

Is your physical activity during leisure-time about as strenuous on average as:

1) walking (corresponding to 4 MET)

2) alternately walking and jogging (6 MET)

3) jogging (10 MET)

4) running (13 MET)

How long does the physical activity last at one session on average:

a) less than 15 minutes (class midpoint 7.5)

b) 15 min – less than 30 min (22.5)

c) 30 min – less than 1 hour (45)

d) 1 hour – less than 2 hours (90)

e) over two hours (120)

Presently how many times per month do you engage in physical activity during your leisure time:

a) less than once a month (class midpoint 0.5)

b) 1-2 times per month (1.5)

c) 3-5 times per month (4)

d) 6-10 times per month (8)

e) 11-19 times per month (15)

f) more than 20 times per month (20)

*Healthy lifestyle*

Lifestyle categories were adapted from Khera et al. (2016).[9] Definitions of the favorable lifestyles used in this study are described in Table S7. A favorable lifestyle included three or four of the healthy lifestyles, intermediate two, and unfavourable one or none.[9]

Table S7. Definitions of the favorable lifestyles

| **Favorable lifestyles** | |
|---|---|
| **No current smoking** | Former smokers or participants who reportedly had never smoked.[9] |
| **No obesity** | Body mass index less than 30 kg · $m^{-2}$.[9] |
| **Physically active** | Meeting the minimum physical activity level according to WHO guidelines.[10] |
| **Healthy diet** | At least six servings of fruits and vegetables per day.[11] |

WHO, World Health Organization.

*Covariates*

Educational attainment describes the self-reported educational attainment by 1981 based on Finnish education system. Low, basic education degree at most; Middle, basic education and additional studies; High, at least upper secondary degree.

Health status is assessed utilizing the most extensive assessment available, including data from Q1981 and data from Finnish nationwide registers between 1971–1983: Nationwide Hospital Discharge Register (NHDR), Social Insurance Institution of Finland (SIIF), and the Finnish Cancer Registry (FCR). 15 Health status is a binary variable, with 1 indicating subject to be ‘healthy’. Subjects with following criteria are defined 0 ‘not healthy’:

- physician-diagnosed angina pectoris, myocardial infarction, or diabetes mellitus (Q1981),
- self-reported history of chest pain (Q1981),
- inpatient admission for diabetes (International Classification of Diseases, Eighth Revision (ICD-8), code 250), cardiovascular disease other than hypertension or venous diseases (ICD-8 codes 390–399 and 410–449), or chronic obstructive pulmonary disease (ICD-8 codes 490–493) between 1972–1982 (NHDR),
- reimbursable medication for selected chronic diseases other than hypertension before January 1, 1983 (SIIF),
- malignant cancer before 1983 (FCR).

Smoking was assessed with multiple questions in 1990 to derive the following categories: Never, no current or prior smoking; Occasional, occasional smoking but never daily; Former, prior regular smoking. Current regular smokers were classified further by smoked cigarettes per day (CPD), Light, 1–9 CPD; Medium, 10–19 CPD; Heavy: > 20 CPD.

Fruit and vegetable consumption was assessed in 1981: “When you are eating a meal or a snack, how many times a day you eat vegetables or fruits (a minimum amount equivalent of one tomato)? Not once, once or twice, three to five times, six time or more.”

Alcohol consumption was based on consumed grams of alcohol per day in 1990.[12] Lifetime abstainer, 0 g and no prior alcohol consumption; Former, 0 g but prior alcohol consumption; Occasional, > 0.1 and < 1.3 g; Low: ≥ 1.3 and < 25 g; Medium: ≥ 25 and < 45 g; High: ≥ 45 g and < 65 g; and Very high: ≥ 65 g per day.

### 3. Supplementary results

#### 3.1 Overall healthy lifestyles

We investigated whether overall healthy lifestyles mitigate the all-cause and cardiovascular mortality in the FTC. The FTC participants were grouped according to their genetic liability for CHD, SBP and DBP and their lifestyles. Reference group was participants with highest genetic disease risk and most unfavourable lifestyles. Analyses were conducted with Cox proportional hazard model. All models were adjusted for ten principal components of ancestry, sex, age, educational attainment, health status, and clustered based on the twin structure of the data. Overall healthy lifestyles were associated with lower risk of all-cause and CVD mortality. Risk reductions were greatest among participants with lowest genetic risk and most favorable lifestyles (HR: 0.24–0.46, Figures S7 and S8).

*All-cause mortality*

A favorable lifestyle was associated with lower risk of mortality across all genetic categories for CHD, but an intermediate lifestyle only in the intermediate genetic risk group. Both favorable and intermediate lifestyles were associated with lower risk of all-cause mortality across all genetic categories for SBP and DBP (Figure S6).

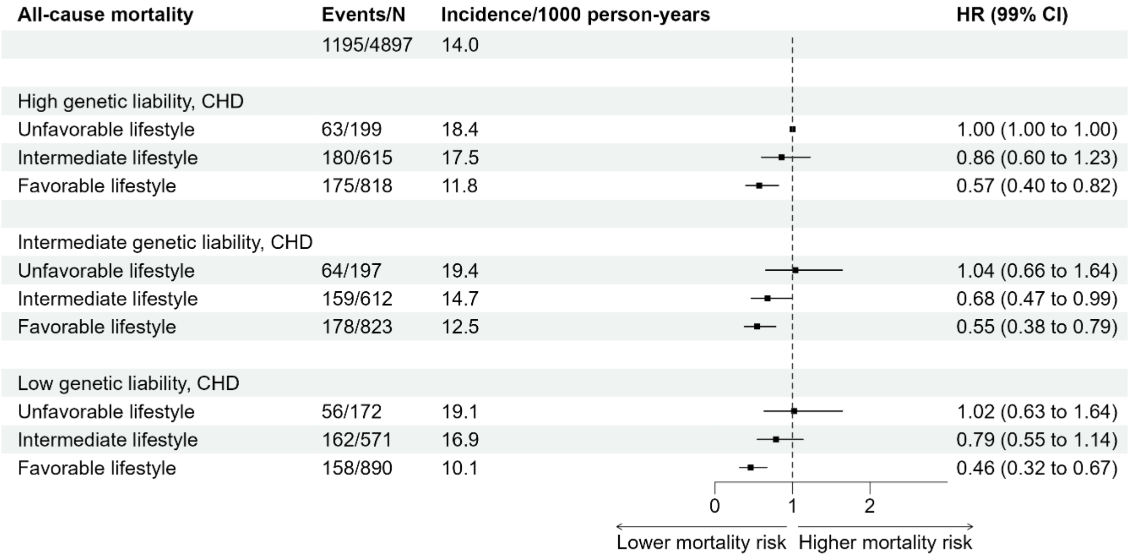

| All-cause mortality | Events/N | Incidence/1000 person-years | HR (99% CI) |
|---|---|---|---|
| | 1195/4897 | 14.0 | |
| High genetic liability, SBP | | | |
| Unfavorable lifestyle | 74/208 | 20.6 | 1.00 (1.00 to 1.00) |
| Intermediate lifestyle | 165/589 | 15.9 | 0.69 (0.48 to 0.98) |
| Favorable lifestyle | 174/835 | 11.6 | 0.45 (0.32 to 0.65) |
| Intermediate genetic liability, SBP | | | |
| Unfavorable lifestyle | 67/202 | 20.1 | 0.84 (0.54 to 1.30) |
| Intermediate lifestyle | 181/608 | 17.4 | 0.65 (0.45 to 0.93) |
| Favorable lifestyle | 174/822 | 12.1 | 0.47 (0.33 to 0.67) |
| Low genetic liability, SBP | | | |
| Unfavorable lifestyle | 42/158 | 15.4 | 0.69 (0.42 to 1.15) |
| Intermediate lifestyle | 155/601 | 15.6 | 0.61 (0.42 to 0.88) |
| Favorable lifestyle | 163/874 | 10.6 | 0.40 (0.28 to 0.58) |

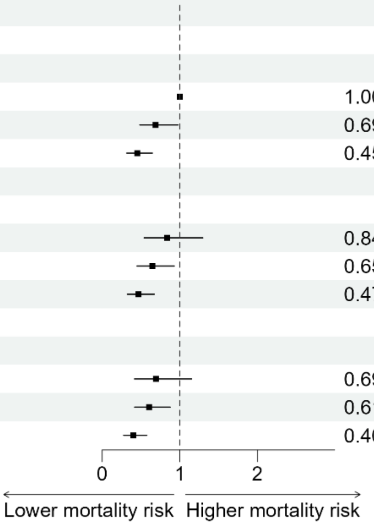

| All-cause mortality | Events/N | Incidence/1000 person-years | HR (99% CI) |
|---|---|---|---|
| | 1195/4897 | 14.0 | |
| High genetic liability, DBP | | | |
| Unfavorable lifestyle | 81/211 | 22.4 | 1.00 (1.00 to 1.00) |
| Intermediate lifestyle | 169/584 | 16.5 | 0.63 (0.45 to 0.89) |
| Favorable lifestyle | 179/837 | 12.0 | 0.45 (0.32 to 0.63) |
| Intermediate genetic liability, DBP | | | |
| Unfavorable lifestyle | 61/190 | 19.5 | 0.86 (0.56 to 1.33) |
| Intermediate lifestyle | 170/629 | 15.8 | 0.61 (0.43 to 0.87) |
| Favorable lifestyle | 162/813 | 11.3 | 0.42 (0.30 to 0.60) |
| Low genetic liability, DBP | | | |
| Unfavorable lifestyle | 41/167 | 14.1 | 0.56 (0.34 to 0.92) |
| Intermediate lifestyle | 162/585 | 16.7 | 0.60 (0.42 to 0.85) |
| Favorable lifestyle | 170/881 | 11.1 | 0.39 (0.27 to 0.55) |

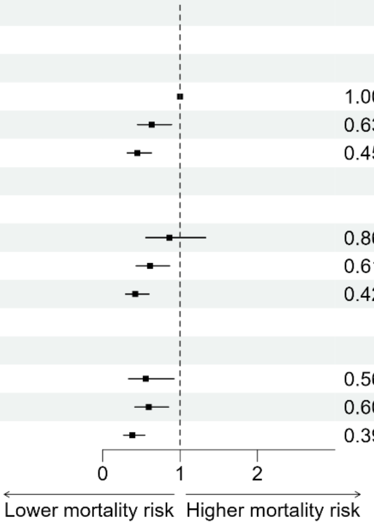

Figure S6. Risk of all-cause mortality (Hazard Ratios and 99% Confidence Intervals) according to different genetic liabilities for cardiovascular diseases and lifestyle categories. Reference group: high genetic liability for disease and unfavourable lifestyle. All models were adjusted for ten principal components, sex, age, educational attainment, health status, and clustered based on the twin structure of the data. CHD, coronary heart disease; SBP, systolic blood pressure; DBP, diastolic blood pressure.

*Cardiovascular mortality*

Favorable lifestyles were associated with lower risk of CVD mortality in all genetic categories for CHD, SBP and DBP, except for high genetic liability for CHD. Intermediate lifestyles were associated with lower risk of CVD mortality across in intermediate and low disease risk categories for SBP and DBP (Figure S7).

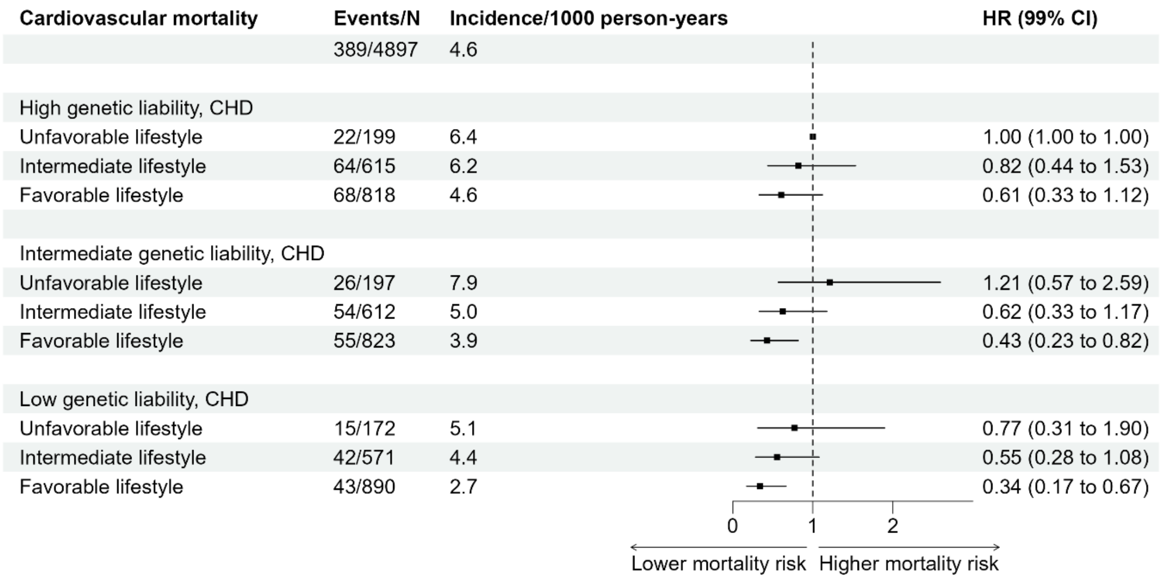

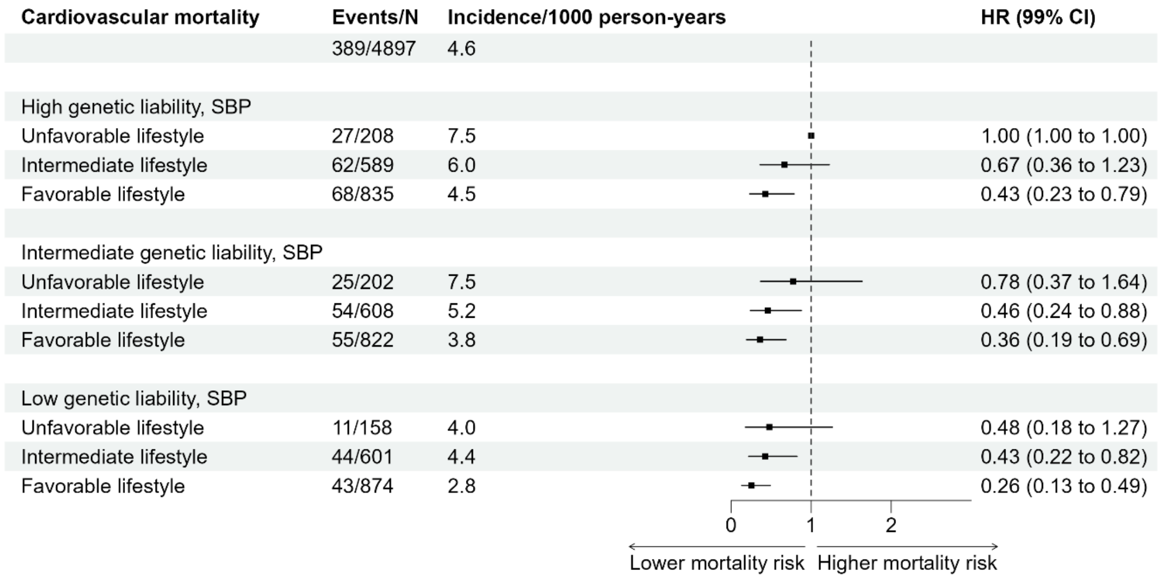

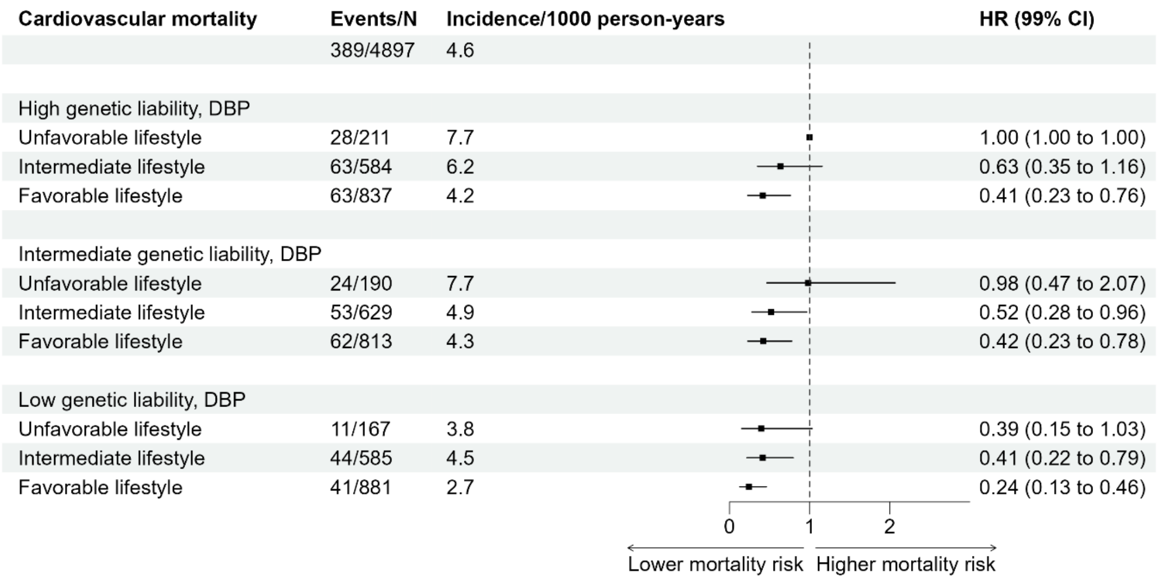

Figure S7. Risk of cardiovascular mortality (Hazard Ratios and 99% Confidence Intervals) according to different genetic liabilities for cardiovascular diseases and lifestyle categories. Reference group: high genetic liability for disease and unfavourable lifestyle. All models were adjusted for ten principal components, sex, age, educational attainment, health status, and clustered based on the twin structure of the data. CHD, coronary heart disease; SBP, systolic blood pressure; DBP, diastolic blood pressure.

##### 3.2 Interactions between genetic liability cardiovascular disease and physical activity

No statistically significant interactions were observed between genetic liability for CVD and different PA variables (Table S8). This finding suggests that physical activity did not moderate the risk of mortality related to inherited liability for CVD. In other words, individuals who had higher genetic predisposition to CVD still had an increased risk of mortality, even if they engaged in physical activity.

Table S8. Genetic disease risk and physical activity interactions with mortality

| Genetic disease risk × Physical activity interaction | 99% Confidence interval | |
|---|---|---|
| | All-cause mortality | CVD mortality |
| **PRS CHD × Meeting PA guidelines (avg MET h · week$^{-1}$ 1975–1990)** | 0.88 to 1.21 | 0.77 to 1.34 |
| **PRS CHD × Meeting PA guidelines 1975, 1981, 1990** | 0.92 to 1.49 | 0.87 to 1.81 |
| **PRS CHD × Vigorous PA 1975, 1981, 1990** | 0.85 to 1.36 | 0.85 to 1.89 |
| **PRS CHD × avg MET (h · week$^{-1}$) 1975–1990** | 0.99 to 1.01 | 0.99 to 1.02 |
| **PRS SBP × Meeting PA guidelines (avg MET h · week$^{-1}$ 1975–1990)** | 0.86 to 1.12 | 0.80 to 1.30 |
| **PRS SBP × Meeting PA guidelines MET 1975, 1981, 1990** | 0.73 to 1.27 | 0.60 to 1.77 |
| **PRS SBP × Vigorous PA 1975, 1981, 1990** | 0.81 to 1.26 | 0.74 to 1.69 |
| **PRS SBP × avg MET (h · week$^{-1}$) 1975–1990** | 0.99 to 1.00 | 0.99 to 1.01 |
| **PRS DBP × Meeting PA guidelines (avg MET h · week$^{-1}$ 1975–1990)** | 0.89 to 1.16 | 0.94 to 1.45 |
| **PRS DBP × Meeting PA guidelines 1975, 1981, 1990** | 0.70 to 1.23 | 0.70 to 1.76 |
| **PRS DBP × Vigorous PA 1975, 1981, 1990** | 0.77 to 1.22 | 0.70 to 1.57 |
| **PRS DBP × avg MET (h · week$^{-1}$) 1975–1990** | 0.99 to 1.00 | 0.99 to 1.01 |

PRSs are Standardised polygenic risk scores. CHD, Coronary heart disease; SBP, systolic blood pressure; DBP, diastolic blood pressure. All models adjusted for ten principal components, sex, age, educational attainment, health status, physical activity, body mass index, smoking, diet, alcohol consumption, and clustered based on the twin structure of the data. Statistically significant interactions (P<0.01) highlighted with **bold.**

Table S9. Associations of polygenic risk scores for systolic blood pressure and physical activity with all-cause mortality after step-by-step adjustments for other healthy lifestyle factors.

| **Risk of all-cause mortality** | **Model 1** | **Model 2** | **Model 3** | **Model 4** | **Model 5** |
|---|---|---|---|---|---|
| **Cases/N** | 1,366 / 5,415 | 1,355 / 5,381 | 1,294 / 5,236 | 1,292 / 5,230 | 1,195 / 4,897 |
| **PRS SBP (ZScore)** | **1.08 (1.00 to 1.15)** | 1.07 (0.99 to 1.15) | 1.05 (0.98 to 1.13) | 1.05 (0.98 to 1.13) | 1.04 (0.96 to 1.12) |
| **Meeting PA guidelines (avg MET h · week$^{-1}$ 1975–1990)** | **0.84 (0.72 to 0.99)** | 0.86 (0.73 to 1.01) | 0.89 (0.76 to 1.05) | 0.91 (0.77 to 1.08) | 0.93 (0.78 to 1.10) |
| **BMI (kg · m$^{-2}$)** | | **1.02 (1.00 to 1.05)** | **1.02 (1.00 to 1.05)** | **1.02 (1.00 to 1.05)** | 1.02 (0.99 to 1.05) |
| **Smoking** | | | | | |
| Never | | | ref | ref | ref |
| Occasional | | | 1.30 (0.85 to 2.00) | 1.30 (0.85 to 2.00) | 1.27 (0.80 to 1.99) |
| Former | | | **1.38 (1.13 to 1.67)** | **1.37 (1.13 to 1.67)** | **1.37 (1.11 to 1.68)** |
| Light | | | **1.80 (1.23 to 2.63)** | **1.79 (1.22 to 2.62)** | **1.75 (1.16 to 2.62)** |
| Medium | | | **2.23 (1.75 to 2.86)** | **2.23 (1.75 to 2.84)** | **2.17 (1.67 to 2.81)** |
| Heavy | | | **3.98 (3.10 to 5.09)** | **3.92 (3.06 to 5.02)** | **3.50 (2.67 to 4.61)** |
| **Fruit and vegetable consumption per day[a]** | | | | | |
| Not once | | | | ref | ref |
| Once or twice | | | | 0.81 (0.65 to 1.01) | 0.87 (0.68 to 1.11) |
| Three times or more | | | | 0.76 (0.53 to 1.08) | 0.81 (0.55 to 1.17) |
| **Alcohol consumption** | | | | | |
| Lifetime abstainer | | | | | ref |
| Former | | | | | 1.02 (0.52 to 2.00) |
| Occasional | | | | | 0.85 (0.58 to 1.25) |
| Low | | | | | 0.94 (0.71 to 1.24) |
| Medium | | | | | 1.31 (0.89 to 1.95) |
| High | | | | | **1.71 (1.01 to 2.91)** |
| Very high | | | | | **2.15 (1.21 to 3.82)** |

Values are Hazard Ratios with 99% Confidence intervals; All models additionally adjusted for ten principal components, age, sex, and educational attainment, health status and clustered based on the twin structure of the data. PRS, polygenic risk score, SBP, systolic blood pressure; ZScore, Standardized score; Meeting PA guidelines, the average of self-reported physical activity from 1975–1990 ≥ 7.5 metabolic equivalent hours per week (MET h · week$^{-1}$); Statistically significant associations (P<0.01) highlighted with **bold.**

Table S10. Associations of polygenic risk scores for systolic blood pressure and physical activity with cardiovascular mortality after step-by-step adjustments with other healthy lifestyle factors.

| **Risk of cardiovascular mortality** | **Model 1** | **Model 2** | **Model 3** | **Model 4** | **Model 5** |
|---|---|---|---|---|---|
| **Cases/N** | 449 / 5,415 | 447 / 5,381 | 426 / 5,236 | 424 / 5,230 | 389 / 4,897 |
| **PRS SBP (ZScore)** | **1.23 (1.08 to 1.40)** | **1.22 (1.07 to 1.39)** | **1.19 (1.04 to 1.36)** | **1.19 (1.04 to 1.37)** | **1.18 (1.03 to 1.36)** |
| **Meeting PA guidelines (avg MET h · week$^{-1}$ 1975–1990)** | 0.77 (0.58 to 1.01) | 0.82 (0.62 to 1.08) | 0.82 (0.62 to 1.10) | 0.86 (0.64 to 1.14) | 0.90 (0.66 to 1.23) |
| **BMI (kg · m$^{-2}$)** | | **1.06 (1.03 to 1.10)** | **1.06 (1.02 to 1.10)** | **1.06 (1.02 to 1.10)** | **1.06 (1.02 to 1.10)** |
| **Smoking** | | | | | |
| Never | | | ref | ref | ref |
| Occasional | | | 1.38 (0.71 to 2.68) | 1.39 (0.72 to 2.71) | 1.36 (0.70 to 2.66) |
| Former | | | 1.32 (0.95 to 1.83) | 1.33 (0.96 to 1.83) | 1.36 (0.96 to 1.94) |
| Light | | | 1.42 (0.70 to 2.89) | 1.42 (0.69 to 2.89) | 1.22 (0.54 to 2.77) |
| Medium | | | **2.10 (1.31 to 3.39)** | **2.12 (1.32 to 3.41)** | **2.10 (1.26 to 3.50)** |
| Heavy | | | **3.68 (2.28 to 5.93)** | **3.60 (2.23 to 5.80)** | **3.35 (1.96 to 5.70)** |
| **Fruit and vegetable consumption per day[a]** | | | | | |
| Not once | | | | ref | ref |
| Once or twice | | | | 0.72 (0.49 to 1.06) | 0.71 (0.47 to 1.08) |
| Three times or more | | | | 0.54 (0.27 to 1.07) | 0.51 (0.25 to 1.05) |
| **Alcohol consumption** | | | | | |
| Lifetime abstainer | | | | | ref |
| Former | | | | | 1.02 (0.39 to 2.65) |
| Occasional | | | | | 0.67 (0.33 to 1.32) |
| Low | | | | | 0.10 (0.51 to 1.30) |
| Medium | | | | | 1.11 (0.55 to 2.25) |
| High | | | | | 1.15 (0.58 to 2.31) |
| Very high | | | | | 1.43 (0.40 to 5.10) |

Values are Hazard Ratios with 99% Confidence intervals; All models additionally adjusted for ten principal components, age, sex, and educational attainment, health status and clustered based on the twin structure of the data. PRS, polygenic risk score, SBP, systolic blood pressure; ZScore, Standardized score; Meeting PA guidelines, the average of self-reported physical activity from 1975–1990 ≥ 7.5 metabolic equivalent hours per week (MET h · week$^{-1}$); Statistically significant associations (P<0.01) highlighted with **bold.**

Table S11. Associations of polygenic risk scores for diastolic blood pressure and physical activity with all-cause mortality after step-by-step adjustments for other healthy lifestyle factors.

| **Risk of all-cause mortality** | **Model 1** | **Model 2** | **Model 3** | **Model 4** | **Model 5** |
|---|---|---|---|---|---|
| **Cases/N** | 1,366 / 5,415 | 1,355 / 5,381 | 1,294 / 5,236 | 1,292 / 5,230 | 1,195 / 4,897 |
| **PRS DBP (ZScore)** | **1.13 (1.05 to 1.22)** | **1.13 (1.05 to 1.21)** | **1.09 (1.02 to 1.18)** | **1.09 (1.01 to 1.18)** | **1.09 (1.01 to 1.18)** |
| **Meeting PA guidelines (avg MET h · week$^{-1}$ 1975–1990)** | **0.85 (0.73 to 0.99)** | 0.86 (0.74 to 1.01) | 0.90 (0.76 to 1.06) | 0.92 (0.78 to 1.08) | 0.93 (0.78 to 1.11) |
| **BMI (kg · m$^{-2}$)** | | **1.02 (1.00 to 1.04)** | **1.02 (1.00 to 1.05)** | **1.02 (1.00 to 1.04)** | 1.02 (0.99 to 1.04) |
| **Smoking** | | | | | |
| Never | | | ref | ref | ref |
| Occasional | | | 1.31 (0.86 to 2.00) | 1.31 (0.85 to 2.00) | 1.27 (0.81 to 1.99) |
| Former | | | **1.37 (1.13 to 1.66)** | **1.37 (1.13 to 1.66)** | **1.36 (1.11 to 1.68)** |
| Light | | | **1.81 (1.23 to 2.64)** | **1.80 (1.23 to 2.63)** | **1.75 (1.17 to 2.34)** |
| Medium | | | **2.23 (1.74 to 2.85)** | **2.22 (1.74 to 2.84)** | **2.15 (1.66 to 2.79)** |
| Heavy | | | **3.94 (3.07 to 5.05)** | **3.88 (3.02 to 4.97)** | **3.46 (2.63 to 4.54)** |
| **Fruit and vegetable consumption per day[a]** | | | | | |
| Not once | | | | ref | ref |
| Once or twice | | | | 0.81 (0.65 to 1.02) | 0.87 (0.68 to 1.11) |
| Three times or more | | | | 0.76 (0.53 to 1.08) | 0.01 (0.55 to 1.17) |
| **Alcohol consumption** | | | | | |
| Lifetime abstainer | | | | | ref |
| Former | | | | | 1.04 (0.53 to 2.03) |
| Occasional | | | | | 0.85 (0.58 to 1.26) |
| Low | | | | | 0.94 (0.71 to 1.25) |
| Medium | | | | | 1.32 (0.89 to 1.96) |
| High | | | | | **1.74 (1.03 to 2.94)** |
| Very high | | | | | **2.21 (1.26 to 3.89)** |

Values are Hazard Ratios with 99% Confidence intervals; All models additionally adjusted for ten principal components, age, sex, and educational attainment, health status and clustered based on the twin structure of the data. PRS, polygenic risk score, DBP, diastolic blood pressure; ZScore, Standardized score; Meeting PA guidelines, the average of self-reported physical activity from 1975–1990 ≥ 7.5 metabolic equivalent hours per week (MET h · week$^{-1}$); Statistically significant associations (P<0.01) highlighted with **bold**

Table S12. Associations of polygenic risk scores for diastolic blood pressure and physical activity with cardiovascular mortality after step-by-step adjustments with other healthy lifestyle factors.

| Risk of cardiovascular mortality | Model 1 | Model 2 | Model 3 | Model 4 | Model 5 |
|---|---|---|---|---|---|
| **Cases/N** | 449 / 5,415 | 447 / 5,381 | 426 / 5,236 | 424 / 5,230 | 389 / 4,897 |
| **PRS DBP (ZScore)** | **1.30 (1.15 to 1.48)** | **1.29 (1.13 to 1.47)** | **1.24 (1.08 to 1.42)** | **1.24 (1.08 to 1.42)** | **1.24 (1.08 to 1.42)** |
| **Meeting PA guidelines (avg MET h · week$^{-1}$ 1975–1990)** | 0.78 (0.59 to 1.04) | 0.83 (0.63 to 1.10) | 0.84 (0.63 to 1.12) | 0.87 (0.65 to 1.17) | 0.92 (0.67 to 1.25) |
| **BMI (kg · m$^{-2}$)** | | **1.06 (1.03 to 1.10)** | **1.06 (1.02 to 1.10)** | **1.06 (1.02 to 1.10)** | **1.06 (1.02 to 1.10)** |
| **Smoking** | | | | | |
| Never | | | ref | ref | ref |
| Occasional | | | 1.40 (0.71 to 2.72) | 1.42 (0.73 to 2.76) | 1.38 (0.70 to 2.70) |
| Former | | | 1.31 (0.95 to 1.82) | 1.32 (0.95 to 1.83) | 1.35 (0.95 to 1.93) |
| Light | | | 1.44 (0.71 to 2.95) | 1.43 (0.70 to 2.94) | 1.25 (0.55 to 2.86) |
| Medium | | | **2.10 (1.31 to 3.38)** | **2.11 (1.32 to 3.39)** | **2.09 (1.26 to 3.48)** |
| Heavy | | | **3.63 (2.25 to 5.85)** | **3.54 (2.19 to 5.71)** | **3.27 (1.91 to 5.60)** |
| **Fruit and vegetable consumption per day[a]** | | | | | |
| Not once | | | | ref | ref |
| Once or twice | | | | 0.73 (0.49 to 1.07) | 0.72 (0.48 to 1.08) |
| Three times or more | | | | 0.54 (0.27 to 1.07) | 0.51 (0.25 to 1.05) |
| **Alcohol consumption** | | | | | |
| Lifetime abstainer | | | | | ref |
| Former | | | | | 1.08 (0.42 to 2.74) |
| Occasional | | | | | 0.68 (0.34 to 1.34) |
| Low | | | | | 0.81 (0.51 to 1.30) |
| Medium | | | | | 1.15 (0.57 to 2.31) |
| High | | | | | 1.19 (0.45 to 3.13) |
| Very high | | | | | 1.51 (0.43 to 5.40) |

Values are Hazard Ratios with 99% Confidence intervals; All models additionally adjusted for ten principal components, age, sex, and educational attainment, health status and clustered based on the twin structure of the data. PRS, polygenic risk score, DBP, diastolic blood pressure; ZScore, Standardized score; Meeting PA guidelines, the average of self-reported physical activity from 1975–1990 ≥ 7.5 metabolic equivalent hours per week (MET h · week$^{-1}$); Statistically significant associations (P<0.01) highlighted with **bold**

##### 3.3 Additional physical activity metrics

Table S13. Independent main effects of different physical activity metrics with all-cause mortality in participants with known genetic liability for coronary heart disease.

| Risk of all-cause mortality | Model 5 |
|---|---|
| **Cases/N** | 1,195/4,897 |
| **avg MET (h · week$^{-1}$) 1975–1990** | 1.00 (0.99 to 1.00) |
| **Cases/N** | 588/2,422 |
| **Meeting PA guidelines 1975, 1981 and 1990** | 0.95 (0.73 to 1.25) |
| **Cases/N** | 543/2,023 |
| **Vigorous PA 1975, 1981 and 1990** | 0.94 (0.72 to 1.23) |

Values are Hazard Ratios with 99% Confidence intervals; Model 5, polygenic risk score for coronary heart disease, ten principal components, sex, age, educational attainment, health status, BMI, smoking, diet and alcohol consumption. Model clustered based on the twin structure of the data. MET h · week$^{1}$, metabolic equivalent hours per week; Statistically significant associations (P<0.01) highlighted with **bold.**

Table S14. Independent main effects of different physical activity metrics with all-cause mortality in participants with known genetic liability for systolic blood pressure.

| Risk of all-cause mortality | Model 5 |
|---|---|
| **Cases/N** | 1,195/4,897 |
| **avg MET (h · week$^{-1}$) 1975–1990** | 1.00 (0.99 to 1.00) |
| **Cases/N** | 588/2,422 |
| **Meeting PA guidelines 1975, 1981 and 1990** | 0.95 (0.73 to 1.24) |
| **Cases/N** | 543/2,023 |
| **Vigorous PA 1975, 1981 and 1990** | 0.94 (0.72 to 1.23) |

Values are Hazard Ratios with 99% Confidence intervals; Model 5, polygenic risk score for systolic blood pressure, ten principal components, sex, age, educational attainment, health status, BMI, smoking, diet and alcohol consumption. Model clustered based on the twin structure of the data. MET h · week$^{1}$, metabolic equivalent hours per week; Statistically significant associations (P<0.01) highlighted with **bold.**

Table S15. Independent main effects of different physical activity metrics with all-cause mortality in participants with known genetic liability for diastolic blood pressure.

| Risk of all-cause mortality | Model 5 |
|---|---|
| **Cases/N** | 1,195/4,897 |
| **avg MET (h · week$^{-1}$) 1975–1990** | 1.00 (0.99 to 1.00) |
| **Cases/N** | 588/2,422 |
| **Meeting PA guidelines 1975, 1981 and 1990** | 0.95 (0.73 to 1.25) |
| **Cases/N** | 543/2,023 |
| **Vigorous PA 1975, 1981 and 1990** | 0.94 (0.72 to 1.23) |

Values are Hazard Ratios with 99% Confidence intervals; Model 5, polygenic risk score for diastolic blood pressure, ten principal components, sex, age, educational attainment, health status, BMI, smoking, diet and alcohol consumption. Model clustered based on the twin structure of the data. MET h · week$^{1}$, metabolic equivalent hours per week; Statistically significant associations (P<0.01) highlighted with **bold.**

Table S16. Independent main effects of different physical activity metrics with cardiovascular mortality in participants with known genetic liability for coronary heart disease.

| Risk of cardiovascular mortality | Model 5 |
|---|---|
| **Cases/N** | 389/4,897 |
| **avg MET (h · week$^{-1}$) 1975–1990** | 1.00 (0.99 to 1.01) |
| **Cases/N** | 201/2,422 |
| **Meeting PA guidelines 1975, 1981 and 1990** | 1.10 (0.68 to 1.78) |
| **Cases/N** | 192/2,023 |
| **Vigorous PA 1975, 1981 and 1990** | 0.97 (0.60 to 1.56) |

Values are Hazard Ratios with 99% Confidence intervals; Model 5, polygenic risk score for coronary heart disease, ten principal components, sex, age, educational attainment, health status, BMI, smoking, diet and alcohol consumption. Model clustered based on the twin structure of the data. MET h · week$^{1}$, metabolic equivalent hours per week; Statistically significant associations (P<0.01) highlighted with **bold.**

Table S17. Independent main effects of different physical activity metrics with cardiovascular mortality in participants with known genetic liability for systolic blood pressure.

| Risk of cardiovascular mortality | Model 5 |
|---|---|
| **Cases/N** | 389/4,897 |
| **avg MET (h · week$^{-1}$) 1975–1990** | 1.00 (0.99 to 1.01) |
| **Cases/N** | 201/2,422 |
| **Meeting PA guidelines 1975, 1981 and 1990** | 1.08 (0.67 to 1.75) |
| **Cases/N** | 192/2,023 |
| **Vigorous PA 1975, 1981 and 1990** | 0.93 (0.58 to 1.49) |

Values are Hazard Ratios with 99% Confidence intervals; Model 5, polygenic risk score for systolic blood pressure, ten principal components, sex, age, educational attainment, health status, BMI, smoking, diet and alcohol consumption. Model clustered based on the twin structure of the data. MET h · week$^{1}$, metabolic equivalent hours per week; Statistically significant associations (P<0.01) highlighted with **bold.**

Table S18. Independent main effects of different physical activity metrics with cardiovascular mortality in participants with known genetic liability for diastolic blood pressure.

| Risk of cardiovascular mortality | Model 5 |
|---|---|
| **Cases/N** | 389/4,897 |
| **avg MET (h · week$^{-1}$) 1975–1990** | 1.00 (0.99 to 1.01) |
| **Cases/N** | 201/2,422 |
| **Meeting PA guidelines 1975, 1981 and 1990** | 1.07 (0.67 to 1.73) |
| **Cases/N** | 192/2,023 |
| **Vigorous PA 1975, 1981 and 1990** | 0.92 (0.58 to 1.48) |

Values are Hazard Ratios with 99% Confidence intervals; Model 5, polygenic risk score for diastolic blood pressure, ten principal components, sex, age, educational attainment, health status, BMI, smoking, diet and alcohol consumption. Model clustered based on the twin structure of the data. MET h · week$^{1}$, metabolic equivalent hours per week; Statistically significant associations (P<0.01) highlighted with **bold.**

##### 3.4 Sensitivity analyses

Sensitivity analyses were conducted to investigate if health status or shorter follow-up time alters the associations. In shorter follow-up design the end of follow-up was set at 15-years earlier (31.12.2005). No statistically significant association were observed (no further data shown). Additionally, analyses were repeated among apparently healthy participants (Table S15 and S16). No statistically significant associations between PA and mortality were observed in individual or pairwise analyses.

*Individual analyses in the apparently healthy population*

Table S19. Independent main effects of different physical activity metrics with all-cause mortality in healthy participants with known genetic liability for coronary heart disease.

| Risk of all-cause mortality | Model 5 |
|---|---|
| **Cases/N** | 927/4,145 |
| **Meeting PA guidelines (avg MET h · week$^{-1}$ 1975–1990)** | 0.97 (0.79 to 1.19) |
| **Cases/N** | 927/4,145 |
| **avg MET (h · week$^{-1}$) 1975–1990** | 1.00 (0.99 to 1.00) |
| **Cases/N** | 477/2,100 |
| **Meeting PA guidelines 1975, 1981 and 1990** | 1.00 (0.74 to 1.35) |
| **Cases/N** | 408/1,705 |
| **Vigorous PA 1975, 1981 and 1990** | 1.00 (0.75 to 1.35) |

Values are Hazard Ratios with 99% Confidence intervals; Model 5, polygenic risk score for coronary heart disease, ten principal components, sex, age, educational attainment, health status, BMI, smoking, diet and alcohol consumption, clustered based on the twin structure of the data. Statistically significant associations (P<0.01) highlighted with **bold.**

Table S20. Independent main effects of different physical activity metrics with all-cause mortality in healthy participants with known genetic liability for systolic blood pressure.

| Risk of all-cause mortality | Model 5 |
|---|---|
| **Cases/N** | 927/4,145 |
| **Meeting PA guidelines (avg MET h · week$^{-1}$ 1975–1990)** | 0.97 (0.80 to 1.19) |
| **Cases/N** | 927/4,145 |
| **avg MET (h · week$^{-1}$) 1975–1990** | 1.00 (0.99 to 1.00) |
| **Cases/N** | 477/2,100 |
| **Meeting PA guidelines 1975, 1981 and 1990** | 1.00 (0.74 to 1.35) |
| **Cases/N** | 408/1,705 |
| **Vigorous PA 1975, 1981 and 1990** | 1.00 (0.74 to 1.34) |

Values are Hazard Ratios with 99% Confidence intervals; Model 5, polygenic risk score for systolic blood pressure, ten principal components, sex, age, educational attainment, health status, BMI, smoking, diet and alcohol consumption, clustered based on the twin structure of the data. Statistically significant associations (P<0.01) highlighted with **bold.**

Table S21. Independent main effects of different physical activity metrics with all-cause mortality in healthy participants with known genetic liability for diastolic blood pressure.

| Risk of all-cause mortality | Model 5 |
|---|---|
| **Cases/N** | 927/4,145 |
| **Meeting PA guidelines (avg MET h · week$^{-1}$ 1975–1990)** | 0.98 (0.80 to 1.20) |
| **Cases/N** | 927/4,145 |
| **avg MET (h · week$^{-1}$) 1975–1990** | 1.00 (0.99 to 1.00) |
| **Cases/N** | 477/2,100 |
| **Meeting PA guidelines 1975, 1981 and 1990** | 1.00 (0.74 to 1.35) |
| **Cases/N** | 408/1,705 |
| **Vigorous PA 1975, 1981 and 1990** | 1.00 (0.74 to 1.34) |

Values are Hazard Ratios with 99% Confidence intervals; Model 5, polygenic risk score for diastolic blood pressure, ten principal components, sex, age, educational attainment, health status, BMI, smoking, diet and alcohol consumption, clustered based on the twin structure of the data. Statistically significant associations (P<0.01) highlighted with **bold.**

*Pairwise analyses for systolic and diastolic blood pressure*

Table S22. Risk of all-cause mortality in more active twins in within-pair comparison with the less active identical twin pair.

| | | Twin meeting PA guidelines |
|---|---|---|
| **Risk of all-cause mortality** | **Cases/N** | **HR (99 % CI)** |
| **By genetic liability for SBP** | | |
| **High** | 28/120 | 0.73 (0.22 to 2.41) |
| **Intermediate** | 28/120 | 0.75 (0.19 to 3.01) |
| **Low** | 31/120 | 0.73 (0.22 to 2.41) |
| **By genetic liability for DBP** | | |
| **High** | 34/120 | 1.34 (0.42 to 4.55) |
| **Intermediate** | 26/120 | 0.67 (0.17 to 2.59) |
| **Low** | 27/120 | 0.46 (0.13 to 1.65) |

Cases, number of deaths; In a co-twin control design with monozygotic twins, twin pairs are controlled for their genetic factors and early life environmental factors. Meeting PA guidelines, the average of self-reported physical activity from 1975–1990 ≥ 7.5 metabolic equivalent hours per week.

Table S23. Risk of cardiovascular mortality in more active twins in within-pair comparison with the less active identical twin pair.

| | | Twin meeting PA guidelines |
|---|---|---|
| **Risk of cardiovascular mortality** | **Cases/N** | **HR (99 % CI)** |
| **By genetic liability for SBP** | | |
| **High** | 14/120 | 0.60 (0.09 to 3.94) |
| **Intermediate** | 10/120 | 0.33 (0.02 to 6.53) |
| **Low** | 12/120 | 0.50 (0.05 to 4.65) |
| **By genetic liability for DBP** | | |
| **High** | 15/120 | 0.75 (0.11 to 5.36) |
| **Intermediate** | 13/120 | 0.29 (0.04 to 2.25) |
| **Low** | 8/120 | 0.33 (0.02 to 6.53) |

Cases, number of deaths; In a co-twin control design with monozygotic twins, twin pairs are controlled for their genetic factors and early life environmental factors. Meeting PA guidelines, the average of self-reported physical activity from 1975–1990 ≥ 7.5 metabolic equivalent hours per week.

*Apparently healthy twins*

Table S24. Risk of all-cause mortality in more active twins in within-pair comparison with the less active identical twin pair among healthy twin pairs.

| | | Twin meeting PA guidelines |
|---|---|---|
| **Risk of all-cause mortality** | **Cases/N** | **HR (99 % CI)** |
| **Among all twin pairs#** | 50/256 | 0.66 (0.26 to 1.67) |
| **By genetic liability for CHD** | | |
| **High** | 22/78 | 0.63 (0.14 to 2.71) |
| **Intermediate** | 13/91 | 0.38 (0.07 to 2.15) |
| **Low** | 15/87 | 1.25 (0.22 to 7.04) |
| **By genetic liability for SBP** | | |
| **High** | 14/84 | 0.67 (0.13 to 3.52) |
| **Intermediate** | 15/85 | 2.00 (0.22 to 18.61) |
| **Low** | 21/87 | 0.33 (0.06 to 1.86) |
| **By genetic liability for DBP** | | |
| **High** | 15/83 | 1.25 (0.22 to 7.04) |
| **Intermediate** | 20/93 | 0.71 (0.16 to 3.23) |
| **Low** | 15/80 | 0.43 (0.07 to 2.55) |

Cases, number of deaths; Excluded twin pairs with at least one twin with known symptoms, diseases, and/or medications. #, Adjusted additionally with BMI, and smoking status; In a co-twin control design with monozygotic twins, twin pairs are controlled for their genetic factors and early life environmental factors. Meeting PA guidelines, the average of self-reported physical activity from 1975–1990 ≥ 7.5 metabolic equivalent hours per week.

Table S25. Risk of cardiovascular mortality in more active twins in within-pair comparison with the less active identical twin pair among healthy twin pairs.

| | | Twin meeting PA guidelines |
|---|---|---|
| **Risk of cardiovascular mortality** | **Cases/N** | **HR (99 % CI)** |
| **Among all twin pairs#** | 21/256 | 0.64 (0.14 to 2.90) |

Cases, number of deaths; Excluded twin pairs with at least one twin with known symptoms, diseases, and/or medications. #, Adjusted additionally with BMI, and smoking status; In a co-twin control design with monozygotic twins, twin pairs are controlled for their genetic factors and early life environmental factors. Meeting PA guidelines, the average of self-reported physical activity from 1975–1990 ≥ 7.5 metabolic equivalent hours per week.

*Pairwise analyses with additional physical activity metrics*

Table S26. Risk of all-cause mortality in more active twins in within-pair comparison with the less active identical twin pair with additional physical activity metrics.

| | | Twin meeting PA guidelines in 1975, 1981 and 1990 |
|---|---|---|
| **Risk of all-cause mortality** | **Cases/N** | **HR (99 % CI)** |
| **Among all twin pairs** | 12/38 | 0.60 (0.14 to 2.51) |

Cases, number of deaths; Excluded twin pairs with at least one twin with known symptoms, diseases, and/or medications. In a co-twin control design with monozygotic twins, twin pairs are controlled for their genetic factors and early life environmental factors. Meeting PA guidelines, self-reported physical activity ≥ 7.5 metabolic equivalent hours per week in each measurement point (1975, 1981 and 1990).

Table S27. Risk of cardiovascular mortality in more active twins in within-pair comparison with the less active identical twin pair with additional physical activity metrics.

| | | Twin meeting PA guidelines in 1975, 1981 and 1990 |
|---|---|---|
| **Risk of cardiovascular mortality** | **Cases/N** | **HR (99 % CI)** |
| Among all twin pairs | 5/38 | 0.50 (0.05 to 5.51) |

Cases, number of deaths; Excluded twin pairs with at least one twin with known symptoms, diseases, and/or medications. In a co-twin control design with monozygotic twins, twin pairs are controlled for their genetic factors and early life environmental factors. Meeting PA guidelines, self-reported physical activity ≥ 7.5 metabolic equivalent hours per week in each measurement point (1975, 1981 and 1990).

Table S28. Risk of all-cause mortality in more active twins in within-pair comparison with the less active identical twin pair with additional physical activity metrics.

| **Risk of all-cause mortality** | **Cases/N** | **Twin with vigorous PA in 1975, 1981 and 1990 HR (99 % CI)** |
|---|---|---|
| Among all twin pairs | 8/36 | 0.75 (0.11 to 5.36) |

Cases, number of deaths; Excluded twin pairs with at least one twin with known symptoms, diseases, and/or medications. In a co-twin control design with monozygotic twins, twin pairs are controlled for their genetic factors and early life environmental factors. Vigorous PA, reported at least intermittent jogging in each measurement point (1975, 1981 and 1990).

Table S29. Risk of cardiovascular mortality in more active twins in within-pair comparison with the less active identical twin pair with additional physical activity metrics.

| **Risk of cardiovascular mortality** | **Cases/N** | **Twin with vigorous PA in 1975, 1981 and 1990 HR (99 % CI)** |
|---|---|---|
| Among all twin pairs | 3/36 | 1.00 (0.03 to 38.20) |

Cases, number of deaths; Excluded twin pairs with at least one twin with known symptoms, diseases, and/or medications. In a co-twin control design with monozygotic twins, twin pairs are controlled for their genetic factors and early life environmental factors. Vigorous PA, reported at least intermittent jogging in each measurement point (1975, 1981 and 1990).

##### 3.4. Exploratory analyses

Associations between genetic disease risk and lifestyle factors were explored with linear regression for continuous variables and multinomial logistic regression for categorical variables. Higher genetic liability for SBP and DBP were associated with higher BMI, but not with other lifestyle factors (Table S24).

Table S30. Associations between genetic liability to cardiovascular disease and different lifestyle factors.

| N=4,897 | Lifestyle factors | | | | | | | |
|---|---|---|---|---|---|---|---|---|
| | **MET 1990 ($h \cdot week^{-1}$)** | **BMI ($kg \cdot m^{-2}$)** | **Smoking** | | **Fruit and vegetable consumption per day** | | **Alcohol consumption** | |
| **CHD Zscore** | -0.44 (-1.22 to 0.36) | 0.12 (-0.02 to 0.26) | never | ref. | not once | ref | lifetime | ref. |
| | | | occasional | 0.17 (-0.05 to 0.39) | 1-2 times | 0.04 (-0.10 to 0.18) | former | 0.06 (-0.36 to 0.48) |
| | | | former | 0.02 (-0.09 to 0.13) | ≥ 3 times | 0.09 (-0.10 to 0.27) | occasional | 0.03 (-0.17 to 0.24) |
| | | | light | 0.01 (-0.17 to 0.18) | | | low | 0.11 (-0.05 to 0.27) |
| | | | medium | 0.14 (-0.01 to 0.28) | | | medium | 0.00 (-0.22 to 0.21) |
| | | | heavy | 0.14 (-0.01 to 0.29) | | | high | 0.05 (-0.25 to 0.36) |
| | | | | | | | very high | -0.08 (-0.44 to 0.29) |
| **SBP ZScore** | -0.25 (-1.00 to 0.51) | **0.24 (0.09 to 0.38)** | never | ref. | not once | ref | lifetime | ref. |
| | | | occasional | -0.00 (-0.22 to 0.21) | 1-2 times | 0.01 (-0.16 to 0.14) | former | 0.14 (-0.27 to 0.55) |
| | | | former | 0.01 (-0.10 to 0.12) | ≥ 3 times | 0.07 (-0.13 to 0.26) | occasional | 0.11 (-0.10 to 0.33) |
| | | | light | 0.09 (-0.08 to 0.27) | | | low | -0.01 (-0.18 to 0.17) |
| | | | medium | 0.07 (-0.08 to 0.21) | | | medium | -0.14 (-0.38 to 0.11) |
| | | | heavy | 0.11 (-0.05 to 0.26) | | | high | -0.12 (-0.47 to 0.23) |
| | | | | | | | very high | -0.10 (-0.57 to 0.36) |
| **DBP ZScore** | 0.06 (-0.70 to 0.82) | **0.27 (0.12 to 0.41)** | never | ref. | not once | ref | lifetime | ref. |
| | | | occasional | -0.03 (-0.26 to 0.20) | 1-2 times | -0.03 (-0.18 to 0.13) | former | 0.11 (-0.29 to 0.52) |
| | | | former | 0.04 (-0.07 to 0.15) | ≥ 3 times | 0.04 (-0.15 to 0.24) | occasional | 0.00 (-0.22 to 0.21) |
| | | | light | 0.03 (-0.15 to 0.21) | | | low | -0.02 (-0.18 to 0.15) |
| | | | medium | 0.08 (-0.06 to 0.23) | | | medium | -0.11 (-0.34 to 0.12) |
| | | | heavy | 0.16 (-0.01 to 0.32) | | | high | -0.12 (-0.46 to 0.23) |
| | | | | | | | very high | -0.22 (-0.64 to 0.20) |

Associations are linear or multinomial logistic regression coefficients per one standard deviation increase in the polygenic risk score and 99% Confidence Intervals. Each model is adjusted for ten principal components, age, sex, educational attainment, health status, and other lifestyle factors. CHD, coronary heart disease; SBP, systolic blood pressure; BDP, diastolic blood pressure; BMI, Body mass index. Statistically significant associations ($P<0.01$) highlighted with **bold.**
