## Supplementary material for "Genetic liability for cardiovascular disease, physical activity, and mortality – findings from The Finnish Twin Cohort": Study Protocol and Statistical Analysis Plan

| | |
|---|---|
| Principal Investigator | Elina Sillanpää, Ph.D<br>Associate Professor<br>Faculty of Sport and Health Sciences, University of Jyväskylä, Finland<br>The Wellbeing services county of Central Finland, Finland |
| Author | Laura Joensuu, Ph.D<br>Postdoctoral Researcher<br>Faculty of Sport and Health Sciences, University of Jyväskylä, Finland |

| | |
|---|---|
| Version | 1.1 |

This Statistical Analysis Plan (SAP) follows the Statistical Analysis Plan guidelines,[1] applied for prospective cohort studies.[2]

#### Signature Page

28.4.2023

Elina Sillanpää, Principal Investigator

Date (DD MM YYYY)

28.4.2023

Laura Joensuu, Author

Date (DD MM YYYY)

#### Abbreviations

| | |
|---|---|
| BMI | Body Mass Index |
| CHD | Coronary Heart Disease |
| CPD | Cigarettes Per Day |
| CVD | Cardiovascular Disease |
| DBP | Diastolic Blood Pressure |
| DNA | Deoxyribonucleic acid |
| FTC | Finnish Twin Cohort |
| ID | Identification Number |
| LTPA | Leisure-time Physical Activity |
| MZ | Monozygotic |
| NA | Not Applicable |
| PA | Physical Activity |
| PRS | Polygenic Risk Score |
| SBP | Systolic Blood Pressure |
| Q1981 | Questionnaire 1981 |
| Q1990 | Questionnaire 1990 |

### 1. Introduction

Previous studies have shown that by adhering to an overall healthy lifestyle (regular physical activity, no obesity, no smoking, and healthy diet) the risk of cardiovascular disease (CVD) mortality could be mitigated despite a risky genotype.[3]

In this study, we focus on the independent role of physical activity (PA) in risk modification and assess whether the risk of all-cause and CVD mortality can be mitigated in individuals despite their inherited CVD risk. PA is commonly observed to be associated with reduced risk of premature death with consistent epidemiological evidence in different demographics with different PA metrics.[4–9]

### 2. Study objectives

#### 2.1 Primary objectives

In this study, our primary objectives are to investigate:

- Does leisure-time PA during adulthood mitigate the risk of all-cause and CVD mortality despite the genetic risk for CVD? We hypothesize that $H_1$: PA is independently associated with reduced risk of mortality despite the genetic CVD risk.
- Are the aforementioned associations causal? We hypothesize that $H_1$: PA is causally associated with reduced risk of all-cause and CVD mortality despite the genetic CVD risk.

#### 2.2 Secondary objectives

Secondary objectives of this study are:

- Is an overall healthy lifestyle associated with reduced risk of all-cause and CVD mortality despite the genetic risk for CVD? We hypothesize that $H_1$: An overall healthy lifestyle is associated with lower risk of all-cause and CVD mortality.

The study utilizes data from the older Finnish Twin Cohort (FTC). First, we assess the secondary objective. We evaluate whether the phenomena where overall healthy lifestyle mitigates risk of mortality replicates in our study sample. Next, we evaluate the primary objectives. In this text, we utilize the terminology of 'individual analyses' with associations between PA and mortality despite the genetic disease risk (analyses of independent main effects). In these analyses the twin cohort is considered as any cohort comprising of individuals. The data twin structure is controlled by standard error clustering. We utilize term 'pair-wise analyses' with assessments related to causality, based on within and between pair comparisons among monozygotic twin pairs. To study potential causal inference between PA and mortality, we utilize a co-twin control design. In this design, pair-wise comparisons in mortality are assessed among monozygotic twins discordant for their physical activity during adulthood. Different metrics of PA are utilized with focus in adherence to World Health Organization's PA guidelines (for aerobic activity). Additional PA metrics include estimates of total volume and intensity.

As sensitivity analyses, the primary objectives are assessed in a subgroup of apparently healthy individuals. For exploratory analyses, the validity of polygenic risk scores (PRSs) as a mortality risk is evaluated, alongside with association between PRSs and healthy lifestyles.

All examined associations are analysed separately with respect to genetic disease risk for coronary heart disease (CHD), systolic blood pressure (SBP), and diastolic blood pressure (DBP).

#### 3. Study Design

We utilize the older FTC described in detail elsewhere.[10] In brief, FTC is a prospective twin cohort and a national resource for genetic epidemiological studies with impressive 45-years of follow-up. All twins identified from the population register of Finland, born before 1958 and living in Finland were contacted in 1975 via questionnaire. A total of 31,145 individuals replied (84 %) and entered the study; this number included persons who satisfied the selection criteria (pairs of persons born on the same day, with the same surname at birth, born in the same local community and of same sex) but who were not twins. Such 'pseudotwins' were not contacted in later surveys. Follow-up questionnaires were sent in 1891 and 1990 to all enrolled and eligible participants.

In this study, we utilize a sub-sample of the FTC who have additionally participated to DNA-sampling in 1993–2017 (N=8,838) (Figure 1). These individuals are genotyped and hence their genetic disease risk level can be assessed. Their vital status in 2020 is inspected based on data from national registers.

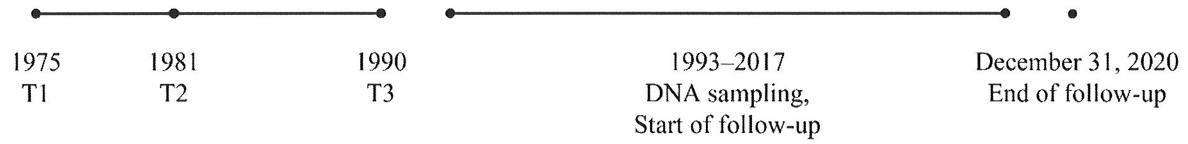

Figure 1. The study design.

##### 3.1 Inclusion and exclusion criteria

The planned inclusion and exclusion criteria with flow of participants for individual analysis and pair-wise analyses are described in Figures 2 and 3.

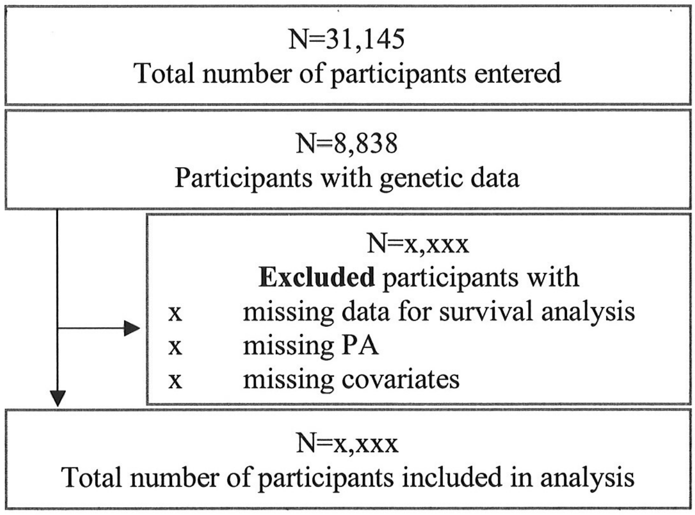

Figure 2. Planned flow of participants through study for individual analyses

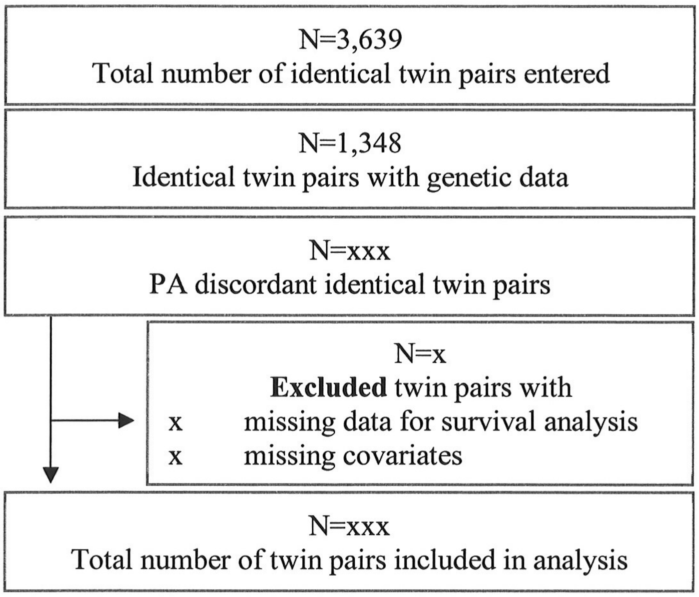

Figure 3. Planned flow of participants through study for pair-wise analyses.

##### 3.2 Sample size and power calculations

Convenience samples are used for analyses. Post-hoc sample size assessments are conducted to estimate the statistical power.

A total of 8,836 participants from the FTC cohort have participated to DNA-sampling. Based on preliminary assessments, 4,897 participants have responded to PA questionnaires at all three measurements points with relevant data for individual analyses. From this sample, a total of 1,195 individuals (24 %) have deceased by the end of 2020. In a recent study, Zhao et al. (2020) found that meeting the current World Health Organization's PA guidelines for aerobic activity was associated with 29 % lower risk of all-cause mortality (Hazard Ratio 0.71).[11] A total of 34% of Finnish adults report to meet the PA guidelines.[12] With 80% power assumption, 34% of subjects in the exposed group, 29% risk reduction in all-cause mortality, and a two-tailed $\alpha$-level of 0.01, a total of 444 death events would be required for the analysis to reach sufficient statistical power.[13]

A co-twin control design is equivalent for a matched case-control study. Informative pairs are the ones discordant for both exposure (in this study PA) and outcome (in this study mortality). For epidemiological studies Odds Ratios (OR) are considered useful for power calculations.[14] Power estimations are conducted post hoc with computer simulations. Based on preliminary assessments, prevalence of mortality in monozygotic twins exposed to inactivity (pE) is 54% and unexposed (pU) 46%. The tetrachoric correlation of the twin-pair in exposure is assessed ($r = 0$). Simulations based on power mcc -analysis are conducted and show that the estimated minimum OR to be detected among the 360 twin pairs identified from the data with 80% power at the .01 significance level is 1.67 (OR), when pU and r are fixed at their observed values. In post hoc analysis, not meeting the PA guidelines had an OR 1.22 (99% CI: 0.76–1.97) against all-cause mortality in the sample. The selected analyzing method Cox regression enables more robust assessment of mortality utilizing the length of the survival time and hence providing more statistical power.

### 4. Covariates

The covariates are based on previous literature. The latest and/or most comprehensive information is utilized. The covariates in individual analyses include ten principal components of ancestry, sex, age, educational attainment, body mass index (BMI), smoking, fruit and vegetable consumption, alcohol consumption and health status. The covariates in pair-wise analyses include the potential main risk factors (BMI and smoking) to adjust for the even minor differences within twin pairs. However, also unadjusted models are used with PA metrics where the number of discordant twin pairs is limited.

*Principal components* of ancestry are continuous variables utilized for correcting potential population stratification in allele frequency.[15] All persons in the cohort are born in Finland.

*Sex* is the register-based variable 'male' or 'female', as documented in birth certification by local parishes and/or local magistrates in the Finnish Population register.

*Age* is whole years since birth to answering the last questionnaire in 1990 (Q1990). Date of birth was recorded in the Central Population register and the date of response to the last questionnaire was recorded upon receipt of the questionnaire.

*Educational attainment* is a categorical variable formed based on the educational structure in Finland (basic education degree at most, basic education and additional studies, or at least upper secondary degree) and indicates the highest degree of education the participant has reported in 1981 questionnaire (Q1981).

*BMI* is square of height in meters ($m^2$) divided by weight (kg). Height and weight were self-reported in Q1990.

*Smoking* is assessed with multiple questions in the Q1990. Smoking is a categorical variable classified as Never = less than 100 cigarettes smoked lifetime, Occasional = occasional smoking but never daily or almost daily, and Former = prior regular (daily or almost daily) smoking. Current regular smokers were classified further as Light = 1–9 smoked cigarettes per day (CPD), Medium = 10–19 CPD, and Heavy: >= 20 CPD.

*Fruit and vegetable consumption* are assessed with Q1981: "When you are eating a meal or a snack, how many times a day you eat vegetables or fruits (a minimum amount equivalent of one tomato)? 0 = not once, 1 = once or twice, 3 = three to five times, 6 = six time of more".

*Alcohol consumption* is assessed in Q1990 and quantified to categories based on consumed grams (g) of alcohol per day, assessed by questions on quantity and frequency of beer, wines and spirits in the past week/month. Lifetime abstainer = 0 g and no prior alcohol consumption, Former = 0 g but prior alcohol consumption, Occasional = 0.1–<1.3 g, Low = 1.3–<25 g, Medium = 25–<45 g, High = 45–<65 g, and Very high = ≥ 65 g per day. Twelve (12) g of alcohol corresponds to a standard drink.

*Health status* is assessed utilizing the most extensive assessment available, including data from Q1981 and data from Finnish nationwide registers between 1971–1983: Nationwide Hospital Discharge Register (NHDR), Social Insurance Institution of Finland (SIIF), and the

Finnish Cancer Registry (FCR).[16] Health status is a binary variable, with 1 indicating subject to be 'healthy'. Subjects with following criteria are defined 0 'not healthy':

- physician-diagnosed angina pectoris, myocardial infarction, or diabetes mellitus (Q1981),
- self-reported history of chest pain (Q1981),
- inpatient admission for diabetes (International Classification of Diseases, Eighth Revision (ICD-8), code 250), cardiovascular disease other than hypertension or venous diseases (ICD-8 codes 390–399 and 410–449), or chronic obstructive pulmonary disease (ICD-8 codes 490–493) between 1972–1982 (NHDR),
- reimbursable medication for selected chronic diseases other than hypertension before January 1, 1983 (SIIF),
- malignant cancer before 1983 (FCR).

#### 5. Study outcomes

##### 5.1 Primary outcomes

Primary outcomes were defined and established a-priori at initiation of the study design. All-cause mortality and CVD mortality are assessed.

All-cause mortality includes all events leading to death. CVD mortality is defined by Finnish national time series classification (54 categories). Categories 27–30 represent CVD mortality. These included ischemic heart diseases (corresponding ICD-10 codes (I20–I25), other forms of heart diseases except rheumatic and alcoholism-based (I30–I425, I427–I52), cerebrovascular diseases (I60–I69), hypertensive, pulmonary heart disease, disease of pulmonary circulation and disease of arteries, arteoles and capillaries, veins, lymphatic vessels, lymph nodes, and other unspecified disorders of the circulatory system (I100–I15, I26–I28, I70–I99).[17,18]

##### 5.2 Secondary outcomes

Lifestyle factors, i.e., BMI, smoking, fruit and vegetable consumption and alcohol consumption.

##### 5.3 Safety outcomes

NA

#### 6. Populations and subgroups of the study

##### 6.1 Study populations

The study population comprised of same-sex twins born before 1958 and living in Finland. In both the FTC and Finnish population approximately 30% of all deaths are CVD related deaths.[19] No differences are observed in the FTC vs. Finnish population in mortality or cancer incidence. Hence, no signs of potential confounding factors related to twin pregnancy (intrauterine growth retardation, low birth weight, or premature birth), i.e. Barker hypothesis with mortality are observed.

##### 6.2 Subgroups

Sensitivity analyses are conducted for a subgroup of apparently healthy subjects as individual's health status might affect their ability to be physically active. The criteria for healthy participants are described in detail in chapter *Covariates*.

### 7. Analyses

##### 7.1 Timing

Interim analyses were performed by Author and preliminary findings presented in relevant academic seminars in 2022. Based on interim analyses, the instruments to assess genetic disease risk, i.e. polygenic risk scores (PRSs) were updated to most recent SBayesR-method, more physical activity variables were adopted to complement the individual and pair-wise analyses, and the analysis method for pair-wise analyses was updated from McNemar's test to more robust Cox proportional hazard model. The final covariates and criteria for sensitivity analyses were selected by the consensus of the research group. Statistical plan was written after accessing the data but prior to conducting the final analysis.

##### 7.2 Handling of missing data and outliers

Potential outliers are inspected with boxplots and whiskers. Due to the length of the observational period, missing data related to physical activity and covariates is expected. A complete-case analysis approach is selected to avoid potential bias caused by imputing data with a twin structure. Missing data analysis are conducted to examine if the data is missing at random.

### 8. Statistical methods

All analyses are conducted with the Stata/IC 16.0 for Windows. Potential moderator effect of sex is tested, and if the interaction term is found non-significant the analyses are conducted combined for both sexes. Interactions between PRSs and PA are tested, and the interaction term is added to the model if statistically significant interaction terms are observed.

##### 8.1 Polygenic risk scores

We utilize Bayesian multiple regression approach and summary statistics from genome-wide association studies to calculate the individual genetic disease risk. The Pan-UK biobank dataset is used as the base data. The Pan-UK Biobank is an open access database with hundreds of thousands of individuals' genetic data paired to electronic health records and survey measures.[20] We select genome-wide summary statistics related CHD, SBP and DBP. and form based on the SBayesR method re-weighted summary statistics restricted to HapMap3 variants.[21,22] The constructed summary statistics include data from 1,005,933–1,006,472 genetic variants and are used to calculate the individual polygenic risk scores for the Finnish Twin Cohort participants. The detailed pipeline is described in detail elsewhere.[23]

The distributions of the PRSs in the FTC are evaluated with a histogram. The PRSs are expected to follow normal distribution. The validity of the PRSs against mortality is tested with crude models (adjusted with ten principal components, and clustered with family number) and Cox proportional hazards model. The PRSs are expected to be associated with higher risk of mortality in the FTC. An increase in risk was calculated per 1 SD change in PRS.

##### 8.2 Descriptive statistics

Characteristics of the individual analyses sample are described by means and standard deviations for continuous variables, and by cases and proportions for binary/categorical variables. The subject descriptives are presented according to vital status in 2020. Similar procedure is conducted for pair-wise analyses.

##### 8.3 Inferential Statistics

**Individual and pair-wise analyses**

Both individual and pair-wise analyses are conducted with Cox proportional hazard model. Cox proportional hazard model does not require data to be normally distributed and can utilize both continuous and binary variables. Cox proportional hazards model rely on two main assumptions: 1) The hazards are proportional over time, and 2) The relationship between log hazards and covariates are linear. These assumptions are tested by Schoenfeld and Martingale residuals.

In all analyses the twin structure of the data is considered: the individual analyses are clustered by the family ID, and the pair-wise analyses stratified by the family ID.

**Examples of related Stata codes**

*Set up and utilities*

All-cause mortality

```
stset exitdate, failure(indic) enter(time entrydate) origin(time birthdate) scale(365.25)
```

CVD mortality

```
stset exitdate, failure(indic_CVD) enter(time entrydate) origin(time birthdate)
scale(365.25)
```

| | |
|---|---|
| exitdate | either date of death, date of emigration, of end of follow-up |
| indic | all-cause mortality yes/no (1/0) |
| indic_CVD | CVD mortality yes/no (1/0) |
| entrydate | date of DNA-sampling |
| birthdate | date of birth |

*Cox proportional hazard model*

Individual analyses

```
xi: stcox ZSCORE_PRS PC1 PC2 PC3 PC4 PC5 PC6 PC7 PC8 PC9 PC10 age i.ea i.sex i.
meetsPAguidelines bmi i.smoking i.fruveg i.alko i.healthy, cl(family_ID) level(99)
```

| | |
|---|---|
| ZSCORE_PRS | standardized score of the polygenic risk score |
| PC1-PC10 | ten principal components of ancestry |
| age | age |
| i.ea | educational attainment (categorical variable) |
| i.sex | sex (male/female) (1/2) |
| i. meetsPAguidelines | meeting physical activity guidelines or not (1/0) |
| bmi | body mass index |
| i.smoking | smoking (categorical variable) |
| i.fruveg | fruit and vegetable consumption (categorical variable) |
| i.alko | alcohol consumption (categorical variable) |
| i.healthy | health status (healthy or not, 1/0) |
| cl(family_ID) | standard errors adjusted for clusters in family_ID |
| level(99) | 99% Confidence Intervals |

Pair-wise analyses

```
xi: stcox i.meetsPAguidelines bmi if MZ_PA_discordantsample==1, strata(family_ID)
level(99)
```

| | |
|---|---|
| i.meetsPAguidelines | meeting physical activity guidelines or not (1/0) |
| bmi | body mass index |
| MZ_PA_discordantsample | analysis conducted within a sample of monozygotic twin pairs discordant for meeting physical activity guidelines |
| strata(family_ID) | data stratified by family_ID for pairwise comparisons |
| level(99) | 99% Confidence Intervals |

**Sensitivity and exploratory analyses**

The sensitivity analyses include individual and pair-wise analyses with similar procedures among apparently healthy subjects only.

Additionally, we evaluate if the PRSs for CVDs derived from the Pan-UK Biobank represent a valid mortality risk in the study population with Cox proportional hazard model. Furthermore, we explore if the genetic disease risk for CVD is associated with healthy lifestyle factors utilizing linear, logistic and multinomial logistic regression. The twin structure of the data is considered by utilizing the svy: regress command.

```
svyset family_ID

svy: regress var1 var2 var3, level(99)
```

**8.4 Statistical significance**

In crude assessments of PRS validity α-level is set to 0.05. In further analysis, a Bonferroni correction is applied with respect to three separate assessments for different genetic disease risk to correct for multiple testing. The α-level is set to a restricted P>0.01 with 99% Confidence Interval.

#### 7. Revision history

| Version/Date | Version name | Section | Changes implemented |
|---|---|---|---|
| Version 1.0/02MAR2023 | Initial draft | NA | NA |
| 1.1/20APR2023 | Final draft | Whole document | Proofreading |

12. *Finland Physical Activity Factsheet*. Europan Comission and WHO Regional Office for Europe https://www.euro.who.int/__data/assets/pdf_file/0008/288107/FINLAND-Physical-Activity-Factsheet.pdf
